# Common variation in meiosis genes shapes human recombination phenotypes and aneuploidy risk

**DOI:** 10.1101/2025.04.02.25325097

**Authors:** Sara A. Carioscia, Arjun Biddanda, Margaret R. Starostik, Xiaona Tang, Eva R. Hoffmann, Zachary P. Demko, Rajiv C. McCoy

**Author notes:** contributed equally (Sara A. Carioscia, Arjun Biddanda).

## Abstract

The leading cause of human pregnancy loss is aneuploidy, often tracing to errors in chromosome segregation during female meiosis ^1,2^. Although abnormal crossover recombination is known to confer risk for aneuploidy ^3–6^, limited data have hindered understanding of the potential shared genetic basis of these key molecular phenotypes. To address this gap, we performed retrospective analysis of preimplantation genetic testing data from 139,416 *in vitro* fertilized embryos from 22,850 sets of biological parents. By tracing transmission of haplotypes, we identified 3,656,198 crossovers, as well as 92,485 aneuploid chromosomes. Counts of crossovers were lower in aneuploid versus euploid embryos, consistent with their role in chromosome pairing and segregation. Our analyses further revealed that a common haplotype spanning the meiotic cohesin *SMC1B* is significantly associated with both crossover count and maternal meiotic aneuploidy, with evidence supporting a non-coding *cis*-regulatory mechanism. Transcriptome- and phenome-wide association tests also implicated variation in the synaptonemal complex component *C14orf39* and crossover-regulating ubiquitin ligases *CCNB1IP1* and *RNF212* in meiotic aneuploidy risk. More broadly, recombination and aneuploidy possess a partially shared genetic basis that also overlaps with reproductive aging traits. Our findings highlight the dual role of recombination in generating genetic diversity, while ensuring meiotic fidelity.

## Introduction

Chromosomes are the physical structures that package DNA, storing genetic information. Despite this crucial role, chromosomes frequently mis-segregate during human meiosis, producing abnormalities in chromosome number—a phenomenon termed "aneuploidy". Aneuploidy is the leading cause of human pregnancy loss, as well as genetic conditions such as Klinefelter, Turner, and Down syndromes ^1,2^. It is estimated that only approximately half of human conceptions survive to birth, primarily due to the abundance of aneuploidies that are inviable in early gestation ^7,8^.

Work in both humans and model organisms has established that one risk factor for aneuploidy involves variation in the number and location of meiotic crossover recombination events, especially in the female germline ^3–6^. Notably, female meiosis initiates in early fetal development, when replicated homologous chromosomes (homologs) pair and establish crossovers, which together with cohesion between sister chromatids hold homologs together in a unique “bivalent” configuration. Homologs segregate (meiosis I) upon ovulation after the onset of puberty, whereas sister chromatids segregate (meiosis II) after fertilization. The physical linkages formed by meiotic crossovers are important for stabilizing paired chromosomes during this prolonged period of female meiotic arrest ^9,10^. Cohesin complexes, loaded in developing fetal oocytes, link sister chromatids and are crucial for chromosome synapsis and crossover formation ^11–13^. Failure to form bivalents due to lack of crossovers ^14,15^ or their suboptimal placement ^16,17^, as well as age-related cohesin deterioration ^18–20^, can lead to premature separation of sister chromatids or reverse segregation (separation of sister chromatids in meiosis I, followed by potentially erroneous separation of homologous chromosomes in meiosis II), which are documented as the predominant mechanisms of maternal meiotic aneuploidy ^20,21^.

While producing high-resolution, sex-specific recombination maps and revealing strong associations with crossover phenotypes at meiosis-related genes such as *PRDM9* and *RNF212*, the largest studies of crossovers in living human families lacked aneuploid individuals and only speculated about such relationships ^22,23^. Much of current knowledge about the connection between human recombination and aneuploidy, as well as their genetic bases, thus comes from smaller samples of living individuals with survivable aneuploidies, limiting statistical power ^24–27^. In contrast, recent advances in single-cell sequencing have enabled simultaneous discovery of crossovers and aneuploidies in sperm and eggs, but are typically relegated to small numbers of gametes (in the case of oocytes) or small numbers of donor individuals, hindering understanding of variance in crossover and aneuploidy phenotypes, as well as their genetic architectures ^28–34^.

Clinical genetic data from preimplantation genetic testing (PGT) of *in vitro* fertilized (IVF) embryos help overcome these limitations and offer an ideal resource for retrospectively characterizing patterns of aneuploidy and mapping meiotic crossovers at scale by comparing haplotypes of multiple sibling embryos ^35^. Here, we used single nucleotide polymorphism (SNP) array-based PGT data from 139,416 blastocyst-stage embryo biopsies and 22,850 sets of biological parents to 1) map recombination and aneuploidy, 2) quantitatively test their relationship, and 3) discover genetic factors that modulate their occurrence and features. Our analysis revealed an overlapping genetic basis of female recombination and aneuploidy formation involving common regulatory variation in key meiotic machinery. Together, our work offers a more complete view of the sources of variation in the fundamental molecular processes that generate genetic diversity while impacting human fertility.

## Results

### Diverse aneuploidies are prevalent in blastocyst-stage embryos and largely trace to errors in maternal meiosis

Seeking insight into meiotic crossover recombination and the origins of aneuploidies, we performed retrospective analysis of clinical genetic data from PGT of human embryos from IVF clinics. Specifically, these data comprised SNP microarray genotyping of bulk (∼6 cells) trophectoderm biopsies from 156,828 blastocyst-stage embryos (5 days post-fertilization), as well as DNA isolated from buccal swabs or blood from both biological parents (24,788 patient-partner pairs; see Methods; **Fig. 1A**, **Fig. S1**, **Fig. S2**). We developed a hidden Markov Model (HMM), called karyoHMM, to trace the transmission of parental haplotypes to sampled embryos and thereby identify aneuploidies and crossover recombination events. Specifically, we modeled transitions between the haplotypes transmitted from the same parent as crossovers and inferred the chromosome copy number that best explained the embryo data (**Fig. 1B**; see Methods) ^36^. The model exhibited high sensitivity and specificity across a range of simulated technical noise parameters that typify data from embryo biopsies (**Fig. S3**).

**Figure 1.**
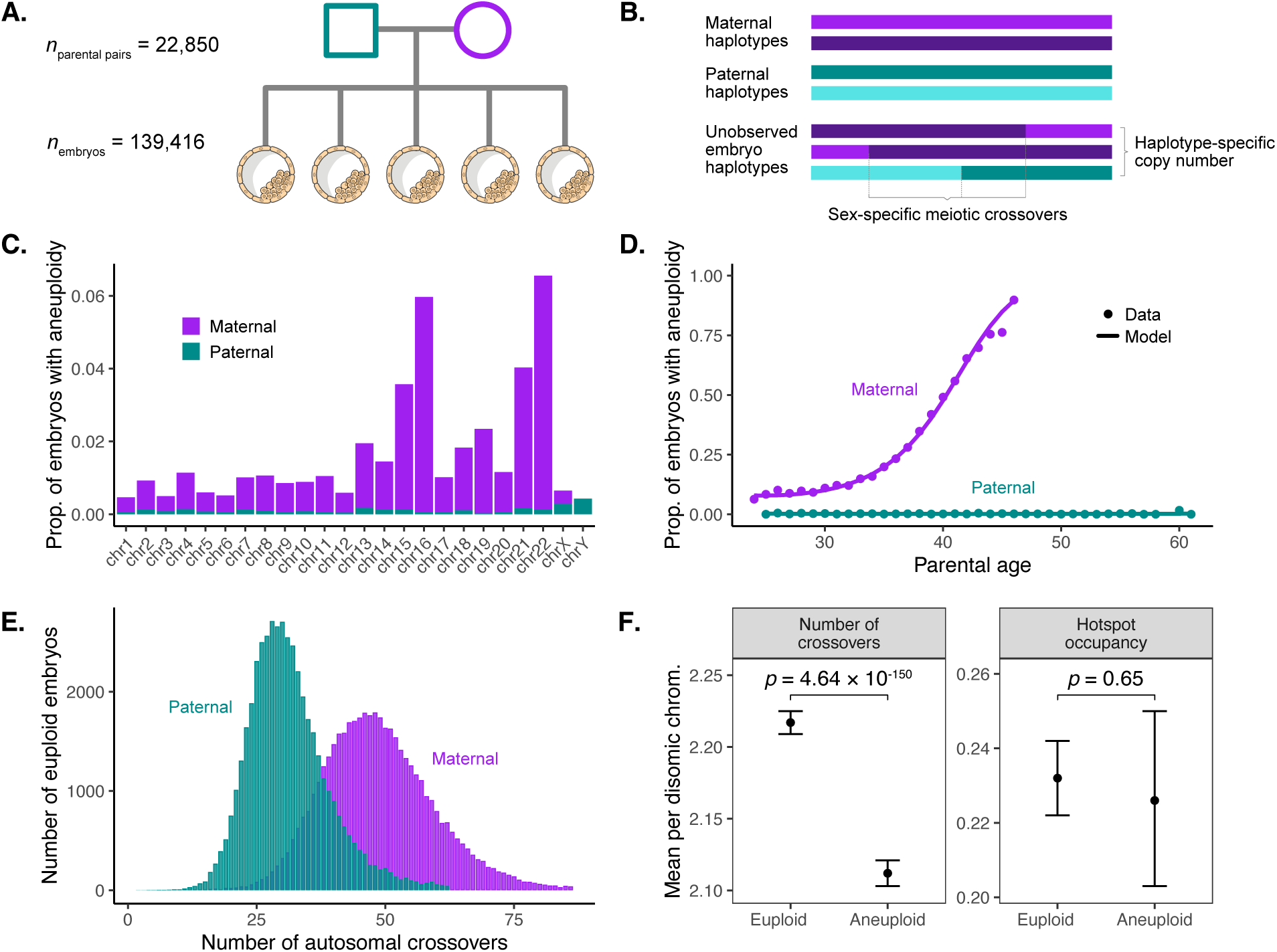
Data from preimplantation genetic testing of IVF embryos offer insight into crossover recombination and aneuploidy. Colors indicate maternal (purple) versus paternal (blue) data features. (A) Data comprise SNP microarray genotyping of trophectoderm biopsies from sibling embryos, as well as DNA from parents. (B) Tracing transmission of parental haplotypes from parents to embryos reveals evidence of crossovers, as well as aneuploidies. (C) Aneuploidies primarily involve gain or loss of maternal homologs and are enriched on particular chromosomes. Complex aneuploidies (>5 affected chromosomes) and genome-wide ploidy abnormalities (e.g., triploidy) are excluded (see **Fig. S4**). (D) Aneuploidies affecting maternal homologs increase with maternal age, while aneuploidies affecting paternal homologs exhibit no significant relationship with paternal age. (E) Maternal crossovers exceed paternal crossovers. Embryos with crossover counts outside of three standard deviations from the sex-specific mean are excluded. (F) Crossover counts differ between disomic chromosomes of aneuploid and euploid embryos, but the proportion of crossovers occurring within hotspots does not.

Applying this method to a filtered dataset where low-quality samples were removed (139,419 remaining embryos; see Methods), we identified 41,480 (29.8%) embryos with at least one aneuploid chromosome (92,485 aneuploid chromosomes; **Fig. S4**). Trisomies exceeded monosomies (57,974 trisomies:34,511 monosomies; ratio = 0.626; binomial test, *p* < 1 × 10^-100^), consistent with selection prior to blastocyst formation ^8^, though trisomies and monosomies of all individual autosomes and sex chromosomes were detected within the sample (**Fig. 1C**). Aneuploidies largely involved gain or loss of maternal versus paternal homologs (84,044 maternal:8,441 paternal; ratio = 0.909; binomial test, *p* < 1 × 10^-100^) and were strongly enriched for chromosomes 15, 16, 21, and 22, consistent with previous literature ^3^^7^. We also replicated the established association between maternal age and the incidence of aneuploidies affecting maternal homologs (binomial generalized linear mixed model (GLMM), *β* = 0.234, *p* < 1 × 10^-100^; **Fig. 1D**) ^20^. The data were well fit by a model with a quadratic term for maternal age, implying that female meiotic aneuploidy accelerates at an approximately constant rate with age (**Table S1**). Despite the statistical power afforded by the large sample size, we observed no significant association between paternal age and aneuploidies affecting paternal homologs (binomial GLMM, *β* = -7.28 × 10^-4^, *p* = 0.956; **Fig. 1D**, **Table S1**), consistent with previous findings ^37^.

### Meiotic crossovers are altered in aneuploid versus euploid embryos

Previous studies have shown that abnormal number or placement of crossovers confers risk for meiotic aneuploidy ^1^. These include studies of survivable trisomies ^25,38^, gametes ^2,31^, and embryos ^30,35^, which broadly demonstrated that aneuploid chromosomes are depleted of crossovers compared to corresponding disomic chromosomes.

Across 46,861 euploid embryos (and requiring ≥ 3 sibling embryos; see Methods), we identified 2,310,257 maternal- and 1,499,155 paternal-origin autosomal crossovers at a median resolution of 99.43 kilobase pairs (kbp) (**Fig. 1E**). The mean counts of sex-specific crossovers per meiosis (49.30 maternal, 31.99 paternal), as well as their genomic locations (Spearman correlation (*r*) at 100 kbp resolution: 0.96 maternal, 0.98 paternal), were consistent with previous pedigree-based studies of living human cohorts ^22,23^. By performing genome-wide association studies (GWAS) across four crossover-derived phenotypes (mean crossover count, hotspot occupancy, replication timing, and GC content; see Methods), we identified 15 unique association signals achieving genome-wide significance (*p* < 5 × 10^-8^), all of which replicated previous findings in the literature ^22^ (**Table S2; Fig. S5 - S8**), including a haplotype spanning *RNF212* with opposing directions of association with maternal versus paternal recombination rates (lead SNP rs3816474; maternal *β* = -0.089, *p* = 1.84 × 10^-11^; paternal *β* = 0.186, *p* = 1.76 × 10^-47^; **Fig. S5**) ^39^. Complementing these GWAS, we also performed transcriptome-wide association studies (TWAS) to associate predicted gene expression across multiple tissues ^40,41^ with recombination phenotypes, identifying 35 unique genes with significant associations with at least one recombination phenotype (*p* < 3.0 × 10^-6^; see Methods; **Table S3**). Prominent hits included the synaptonemal complex component *C14orf39* (also known as *SIX6OS1*) ^42^ and crossover-regulating ubiquitin ligase *CCNB1IP1* (also known as *HEI10*) ^43^, implying that previously reported genetic associations at these loci could be driven by non-coding regulatory mechanisms ^22^.

To examine the relationship between crossovers and aneuploidies, we contrasted patterns of crossovers between aneuploid and euploid embryos within our dataset. One technical limitation for direct detection of crossovers using genetic data from trisomic chromosomes is that crossovers can be missed when both reciprocal products of a single crossover event are transmitted to the embryo ^30^. To overcome this concern, we instead contrasted counts of crossovers on disomic chromosomes of aneuploid embryos (where the aneuploidy affected a different chromosome) to corresponding disomic chromosomes of euploid embryos. This comparison relies on the previous observation that crossovers exhibit positive covariance across chromosomes within a given meiocyte ^44^—a phenomenon that we replicate for euploid embryos within our dataset (intraclass correlation coefficient (ICC) = 0.176, *p* < 1 × 10^-100^ maternal; 0.088, *p* < 1 × 10^-100^ paternal; see Methods; **Fig. S9**). As input to our test, we identified 1,914,536 maternal and 1,290,261 paternal-origin crossovers on disomic chromosomes across 43,577 embryos with at least one chromosome inferred to be aneuploid. Using a Poisson GLMM (see Methods), we found that the number of crossovers was significantly lower on the disomic chromosomes of aneuploid embryos relative to euploid embryos (*β* = 0.105 difference in marginal means, *p* = 1 × 10^-306^, **Fig. 1F**). These results are consistent with the understanding that reduction in crossovers—and absence of crossovers, in particular ^45,46^—is a key risk factor in the origins of aneuploidies.

### A common haplotype spanning the meiotic cohesin *SMC1B* is associated with maternal meiotic aneuploidy

Previous studies have suggested that the incidence of female meiotic aneuploidy may be individual-specific, even after accounting for the known effect of maternal age ^47,48^. To test this hypothesis in our data, we fit a quasi-binomial generalized linear model (GLM) to the per-patient counts of embryos affected versus unaffected with maternal meiotic-origin aneuploidy, including maternal age as a quadratic covariate (see Methods). We then simulated new counts of affected and unaffected embryos from the fitted model for the same size sample, but assuming no overdispersion (i.e., binomial; *n* = 1,000 replicate simulations). Compared to this simulated null distribution, the observed incidence of meiotic aneuploidy was significantly overdispersed across female individuals, controlling for maternal age (dispersion parameter (*φ*) = 1.15, *p* < 0.01; **Fig. S10**). These results are consistent with a role of genetic and environmental factors beyond age in observed variation in maternal meiotic aneuploidy.

To investigate the genetic component, we scanned for variation in maternal genomes associated with the incidence of maternal meiotic aneuploidy. We implemented this GWAS using a binomial GLMM, controlling for covariates including maternal age (see Methods). Our analysis revealed two associations achieving genome-wide significance (*p* < 5 × 10^-8^) (**Fig. 2A, Fig. S11**). The first hit (lead SNP rs9351349, *β* = 0.078, *p* = 2.93 × 10^-8^) lies within an intergenic region of chromosome 6 but did not replicate in a held-out test set comprising 15% of female individuals (*β* = 0.021, *p* = 0.529). The second hit (lead SNP rs6006737, *β* = 0.066, *p* = 2.21 × 10^-8^) lies on chromosome 22 and replicated in the held-out test set (*β* = 0.059, *p* = 0.033). The minor (C) allele within our sample is globally common, segregating at high frequencies (gnomAD AF = 0.78) in African populations but at lower frequencies in European (gnomAD AF = 0.35) and other non-African populations ^49^. The effect is additive, whereby for a 40-year-old patient, each copy of the risk allele confers an estimated 1.65% additional average risk of aneuploidy (**Fig. 2B**). We also detected evidence of a small but statistically significant interaction between maternal age and genotype (Likelihood ratio test, *χ*^2^ (1) = 4.24, *p* = 0.040), indicating that the effect of genotype increases with increasing maternal age (*β* = 0.026, *p* = 0.045). Notably, the size and direction of the main effect of genotype is relatively consistent for aneuploidies of all individual autosomes (**Fig. S12**), suggesting broad, genome-wide impacts on meiotic fidelity.

**Figure 2.**
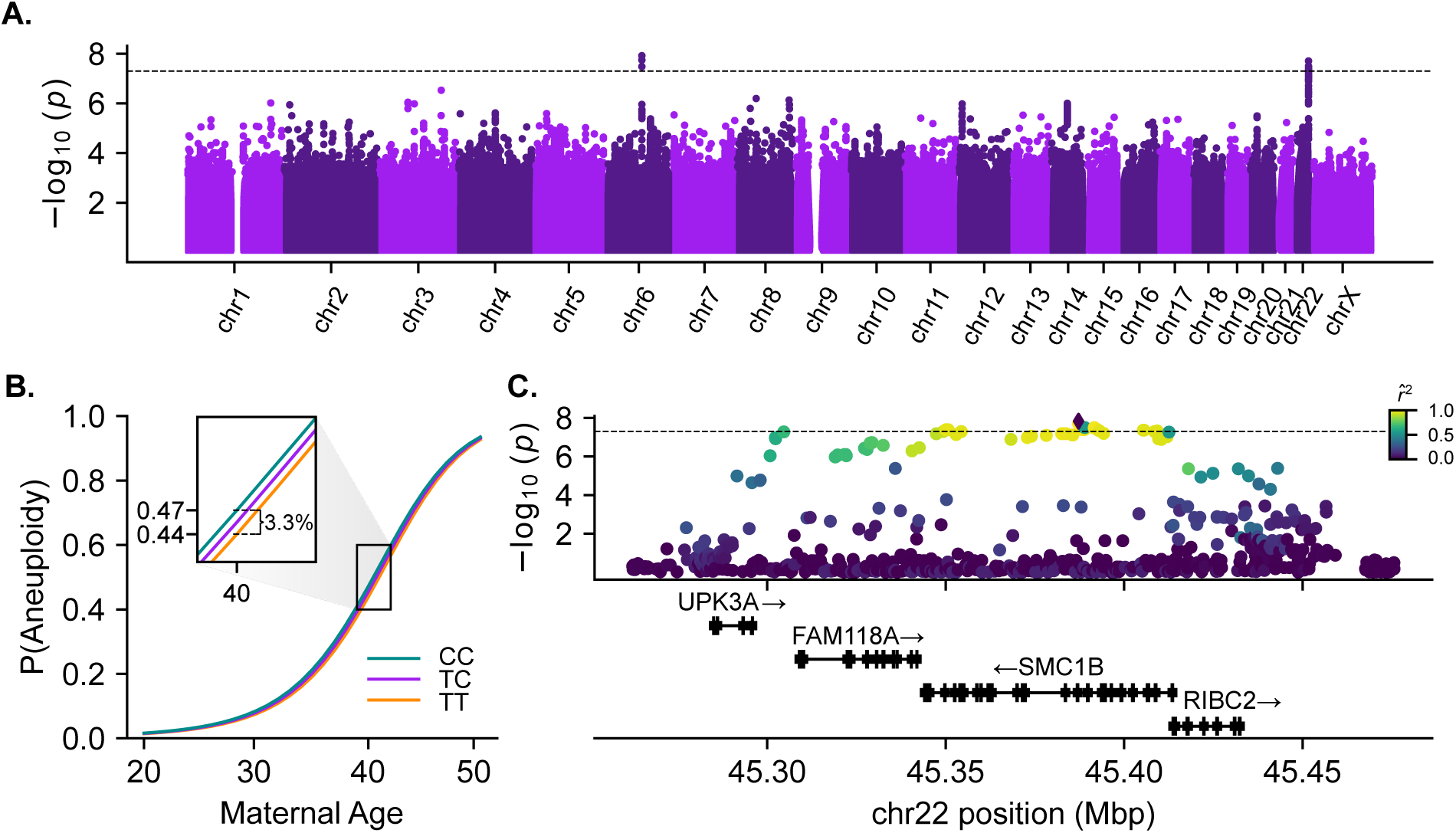
Variants defining a haplotype spanning *SMC1B* are associated with incidence of maternal meiotic aneuploidy. (A) Genome-wide association tests of maternal meiotic aneuploidy and maternal genotype. The dotted line indicates the threshold for genome-wide significance (*p* = 5 x 10^-8^). (B) Fitted relationship between maternal age and incidence of aneuploidy, stratified by maternal genotype at aneuploidy-associated lead SNP rs6006737. (C) Regional association plot depicting the associated locus on chromosome 22, with points colored based on pairwise linkage disequilibrium with the lead SNP rs6006737.

The associated haplotype spans approximately 120 kbp, encompassing four genes: *UPK3A*, *FAM118A*, *RIBC2*, and *SMC1B* (**Fig. 2C**). *SMC1B* encodes a component of the ring-shaped cohesin complex (**Fig. 3A**), with integral roles in sister chromatid cohesion and homologous recombination during meiosis ^50–52^. *SMC1B*-deficient mice of both sexes are sterile, and females exhibit meiotic abnormalities including reduction in crossovers, incomplete chromosome synapsis, as well as age-related premature loss of sister chromatid cohesion and chromosome mis-segregation ^50,51^. Previous work in humans has demonstrated associations between a less common (gnomAD global AF = 0.06) *SMC1B* missense variant (rs61735519; *r*^2^ with GWAS lead SNP rs6006737 = 0.089, *D’* = 0.943) and recombination phenotypes ^22^. Although imputed with moderate accuracy (dosage *r*^2^ = 0.80), this missense variant exhibits only modest association with aneuploidy within our sample (*β* = 0.112, *p* = 4.80 × 10^-3^). Meanwhile, the more common aneuploidy-associated haplotype tagged by GWAS lead variant rs6006737 lacks amino acid altering variation (*r*^2^ < 0.1 for all *SMC1B* nonsynonymous variants), motivating us to explore potential regulatory mechanisms driving the observed phenotype.

**Figure 3.**
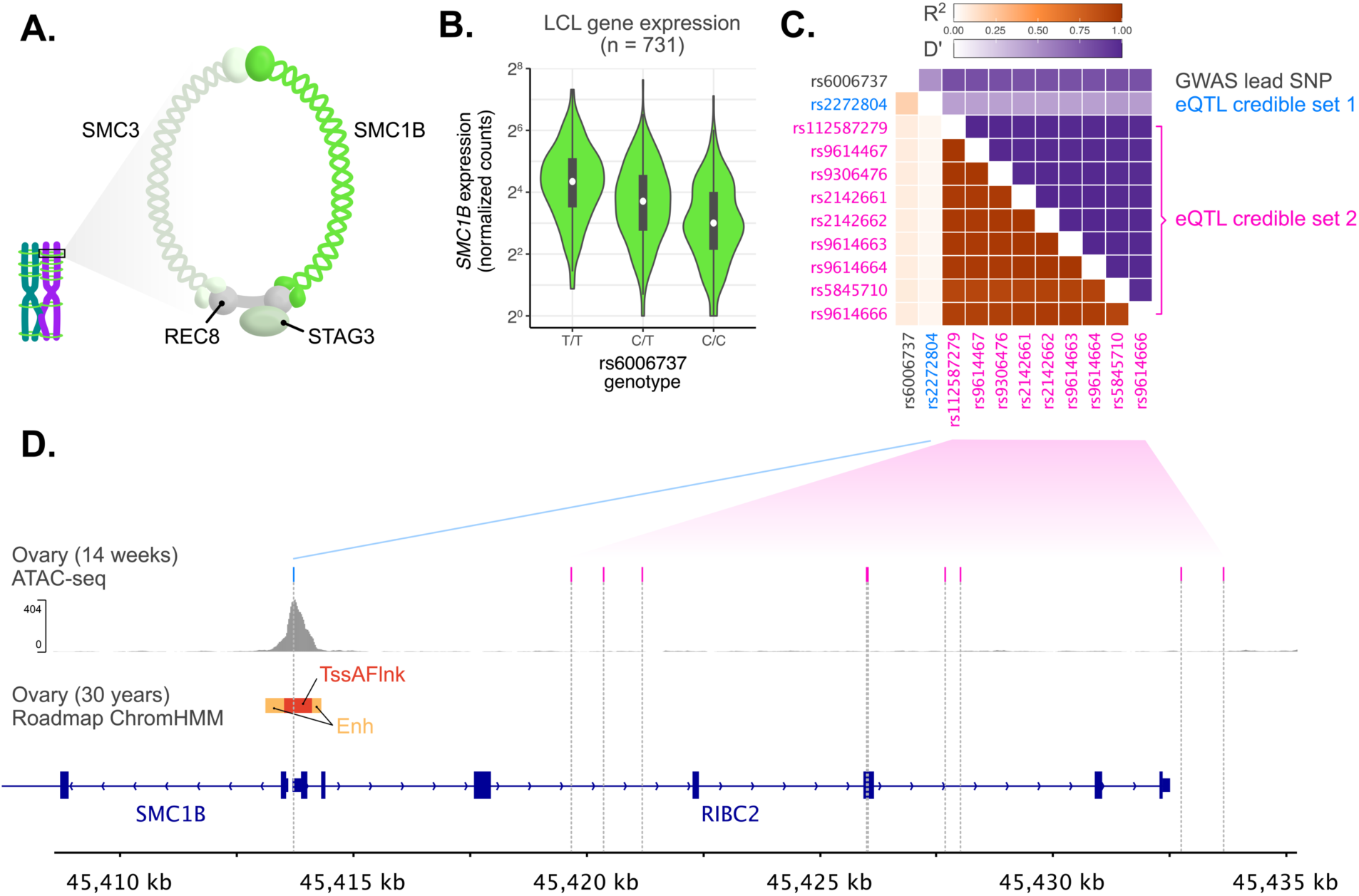
The aneuploidy risk haplotype is associated with lower expression of *SMC1B*, driven by two independent causal signals. (A) Schematic of the meiotic cohesin complex. (B) Each copy of the aneuploidy risk allele is associated with reduced expression of *SMC1B* in cell lines from diverse human populations. (C) Pairwise linkage disequilibrium between a set of SNPs including GWAS lead SNP rs6006737 and variants defining fine-mapped eQTL credible sets for *SMC1B*. (D) Fine-mapped eQTL rs2272804 (credible set 1) lies within a putative promoter sequence within open chromatin, while variants defining a second credible set are distributed throughout the upstream region of *SMC1B*.

### The aneuploidy risk allele is associated with reduced expression of *SMC1B*

Querying the GWAS lead variant (rs6006737) in data from the Genotype Tissue Expression (GTEx) Project, we observed that the aneuploidy risk allele is significantly associated with reduced expression of *SMC1B* across diverse tissues ^41^. While invaluable, GTEx largely includes subjects of European ancestries, limiting resolution for fine-mapping of causal expression-altering variants. To address this limitation, we also queried the GWAS lead variant in MAGE, which includes RNA-seq data from lymphoblastoid cell lines from 731 individuals from 26 globally diverse populations ^53^. Consistent with GTEx, rs6006737 is a strong eQTL of *SMC1B* in MAGE (*β* = -0.429, *p* = 4.68 × 10^-18^; **Fig. 3B**). Fine-mapping within MAGE decomposes the eQTL signals for *SMC1B* into two credible sets containing candidate causal variants (coverage = 0.95) (**Fig. 3C** & **3D**). While one credible set includes nine variants distributed throughout the upstream region of *SMC1B*, the other credible set is defined by a single SNP (rs2272804; posterior inclusion probability (PIP) > 0.99), 144 bp upstream of the *SMC1B* transcription start site. The (A) allele of rs2272804 associated with lower *SMC1B* expression (and higher aneuploidy risk) is globally common (gnomAD global AF = 0.44), with higher frequencies among African populations (gnomAD AF = 0.71). While the putative ancestral (C) allele appears fixed among extant non-human great ape populations ^54^, the variant is polymorphic across high-coverage Neanderthal genomes ^55,56^, and coalescence-based methods estimate that the derived allele originated 910,650 years ago (95% CI: 825,825-1,004,175) ^57^.

The regulatory potential and accessibility of the putative promoter CpG island sequence within which rs2272804 resides is supported by published epigenomic and ATAC-seq data from human ovaries ^58,59^ (**Fig. 3D**). We further noted that the SNP lies within a predicted binding motif of ATF1 ^60^, a transcription factor expressed in female germ cells ^61^ and previously inferred to regulate paralog *SMC1A* based on ChIP-seq data ^62^. Binding of ATF1 to the SNP-encompassing locus is additionally supported by high-confidence ChIP-seq peaks in induced pluripotent stem cells (WTC11) assayed by the ENCODE Project ^62^. By performing an electrophoretic mobility shift assay (EMSA), we found that a DNA construct containing the alternative allele of rs2272804 had more than three-fold lower binding affinity for purified human ATF1 *in vitro* than a construct containing the reference allele (Student’s *t*-test, mean reference K_D_ = 56.62 nM ± 4.65 SD, mean variant K_D_ = 173.39 nM ± 15.24 SD, *p* = 2.60 × 10^-4^), consistent with the observed eQTL effect (**Fig. S13**). Taken together, these results suggest a potential non-coding regulatory mechanism underlying the observed genetic association with maternal meiotic aneuploidy.

### *Cis*-regulatory effects on expression of additional meiosis-related genes are further associated with aneuploidy risk

Motivated by our observations at *SMC1B*, we next sought to examine whether other *cis*-regulatory effects on expression could influence aneuploidy risk. To this end, we again used TWAS to test whether predicted gene expression across tissues is associated with incidence of aneuploidy (see Methods). Across 16,685 protein-coding genes, we identified two hits achieving transcriptome-wide significance (*p* < 3 × 10^-6^; **Fig. 4A**). Although led by adjacent gene *RIBC2* (*p* = 2.19 × 10^-7^), the peak on chromosome 22 includes *SMC1B* (*p* = 7.63 × 10^-6^), replicating our findings from GWAS and downstream functional dissection. We hypothesize that *RIBC2* represents a secondary, noncausal association, whereby the same haplotype (and potentially the same causal variant ^63^) co-regulates expression of both genes, driving their correlation (**Fig. S14**). The second peak on chromosome 14 is led by *C14orf39* (*p* = 1.65 × 10^-7^), which encodes a component of the central element of the synaptonemal complex—the evolutionarily conserved zipper-like structure that mediates synapsis, recombination, and segregation of homologous chromosomes during meiosis ^64^. Previous studies have linked rare *C14orf39* variants to human infertility ^65–67^ and demonstrated associations between common *C14orf39* variants and recombination phenotypes ^22,68^. Our results connect these findings and show that both rare and common variation influencing female fertility differences can converge on the same meiosis-related genes. While not achieving transcriptome-wide significance, a third peak on chromosome 12 includes *NCAPD2* (*p* = 2.16 × 10^-5^), which encodes a regulatory subunit of the condensin I complex, involved in chromosome condensation during both meiotic and mitotic prophase ^69^. Together, our findings highlight the role of common non-coding *cis*-regulatory variation influencing expression of meiosis-related genes in modulating risk of maternal meiotic aneuploidy (**Fig. 4B**).

**Figure 4.**
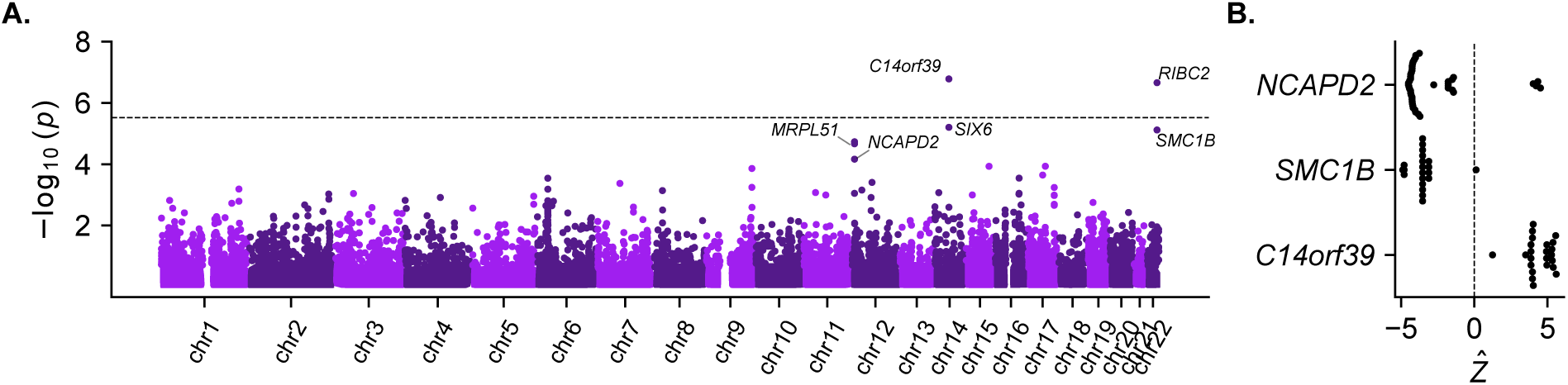
Transcriptome-wide association study (TWAS) for maternal meiotic aneuploidy. (A) Transcriptome-wide association tests of maternal meiotic aneuploidy and predicted maternal gene expression, combining across tissues (see Methods). The dotted line indicates the threshold for transcriptome-wide significance (*p* = 3 × 10^-6^). (B) Per-tissue Z-scores indicating the direction of association between predicted expression and maternal meiotic aneuploidy.

### A shared genetic basis of recombination, aneuploidy, and other fertility-related traits

Given the relationship between crossovers and aneuploidies, we next sought to contextualize our association findings and examine the potential shared genetic basis of these phenotypes and other fertility-related traits. To this end, we identified the lead variant from each genome-wide significant peak in female recombination and aneuploidy GWAS and queried their associations with all recombination and aneuploidy phenotypes, as well as published GWAS of female reproductive aging and infertility traits (i.e., phenome-wide association). Our analysis revealed that the risk allele of the aneuploidy-associated lead SNP rs6006737 is also associated with lower rates of female recombination within our data (*β* = -0.033, *p* = 0.002), consistent with the known role of *SMC1B* variation in this phenotype ^50^. Extending to published GWAS data ^70,71^, we observed that the aneuploidy risk allele is additionally associated with greater age at menarche (*β* = 0.021, *p* = 3.82 × 10^-12^) and lesser age at menopause (*β* = -0.047, *p* = 2.06 × 10^-4^) and thus a shorter female reproductive timespan (**Fig. 5**).

**Figure 5.**
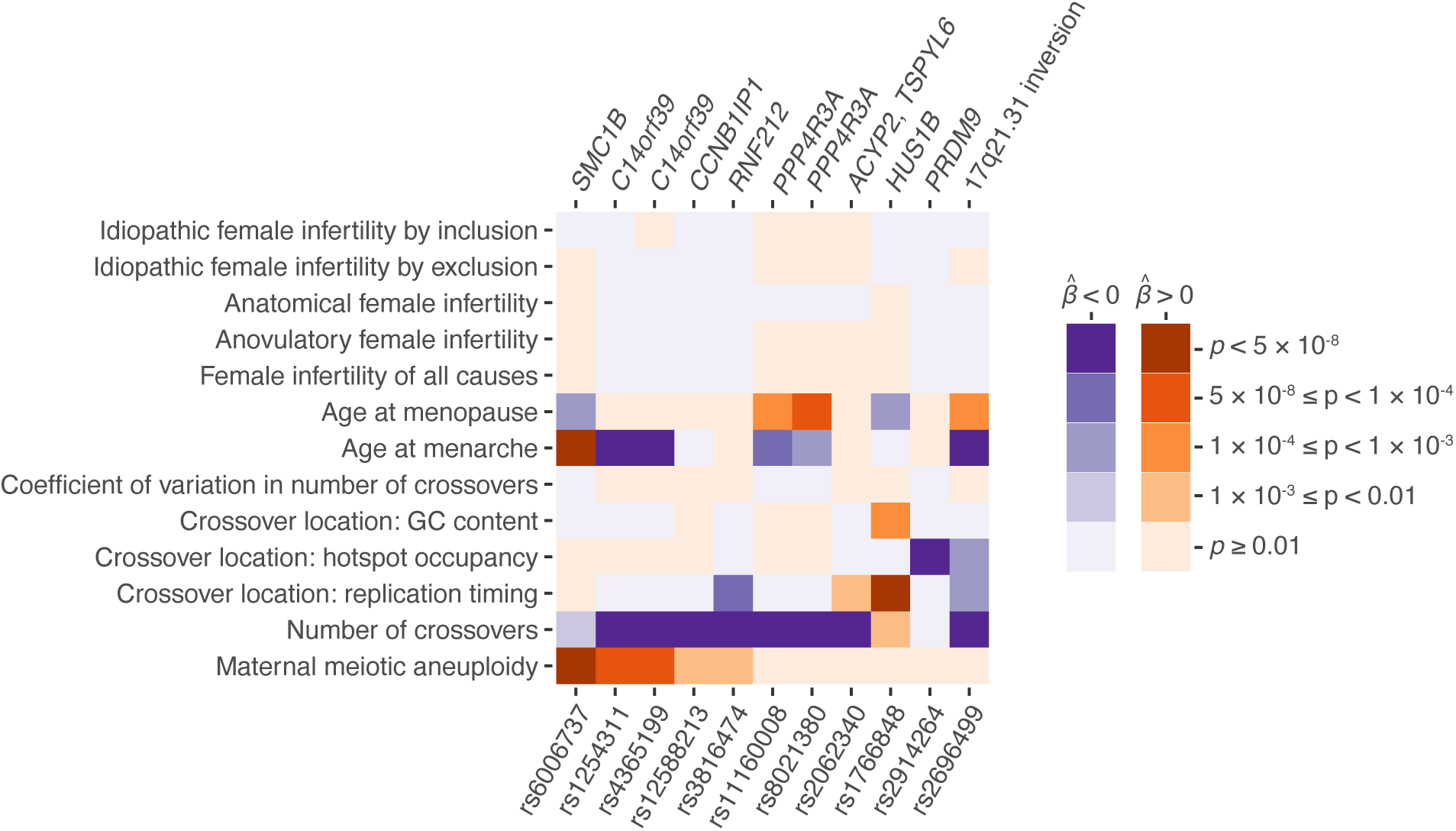
Aneuploidy, recombination, and female reproductive aging traits share an overlapping genetic basis. The lead SNP from each peak from GWAS of aneuploidy and recombination was queried for association with other fertility-related phenotypes. Darkness indicates significance of association (*p*-value), while color indicates direction of association. SNPs are polarized such that the aneuploidy-increasing allele is queried across all traits. Each hit is labeled based on meiosis-related candidate genes within the associated region (top), with the exception of the common 17q21.31 inversion, as well as the locus containing *ACYP2* and *TSPYL6*, where no such candidate is apparent.

Strikingly, we also found that three of the genome-wide significant hits for female recombination rate (**Table S2**) exhibited nominal associations with aneuploidy. In all such cases, the allele associated with a lower rate of recombination was associated with a higher rate of aneuploidy. The first hit (lead SNP rs4365199; aneuploidy *β* = 0.056, *p* = 5.58 × 10^-6^; gnomAD global AF = 0.39) comprises a 175 kbp haplotype spanning synaptonemal complex component *C14orf39*, consistent with our previous TWAS results. The second hit (lead SNP rs12588213; aneuploidy *β* = 0.037, *p* = 1.46 × 10^-3^; gnomAD global AF = 0.42) comprises a 15 kbp haplotype spanning *CCNB1IP1*, encoding an E3 ubiquitin ligase demonstrated as essential for crossover maturation and fertility in mice ^72^. The last hit (lead SNP rs3816474; aneuploidy *β* = 0.041, *p* = 5.04 × 10^-3^; gnomAD global AF = 0.22) comprises a 59 kbp haplotype spanning the E3 ubiquitin ligase *RNF212*, encoding an essential regulator of meiotic recombination that interacts with CCNB1IP1 and helps to designate sites of crossovers versus non-crossovers ^73^. Several of these recombination and aneuploidy-associated variants also exhibited secondary associations with ages at menarche and menopause (**Fig. 5**). While previous studies have reported pleiotropic effects whereby variants that disrupt DNA damage repair are associated with higher rates of *de novo* point mutations and earlier age at menopause ^70,74–76^, the inconsistencies in directions of effects in our data imply that the relationship with aneuploidy may be more complex. Moreover, none of the aneuploidy-associated variants exhibited even nominal associations with various definitions of female infertility ^77^, potentially reflecting the multifactorial nature of clinical infertility.

Despite our discoveries of several genome- and transcriptome-wide significant loci, the proportion of variance in maternal meiotic aneuploidy explained by genotyped SNPs (i.e., SNP heritability) was negligible (*h*^2^_SNP_ = 0.023 ± 0.024 SE; **Table S4**), though SNP heritability of female recombination rate was moderately higher (*h*^2^_SNP_ = 0.112 ± 0.042). These estimates are in line with low reported SNP heritabilities of female fertility phenotypes ^77^ and consistent with evolutionary theory and previous observations regarding other fitness-related traits ^78,79^. Given these observations, we hypothesized that environmental factors ^80^ and rare genetic variation likely contribute to residual variance in aneuploidy rates, including effects on meiotic recombination. In support of this hypothesis, individual-specific rates of recombination were inversely associated with aneuploidy, even after controlling for maternal age and all aforementioned genetic associations (binomial GLMM, *β* = -0.763, *p* = 8.15 × 10^-8^; see Methods). The direction of association, whereby lower rates of recombination are associated with higher rates of aneuploidy, is consistent with our reported embryo-level patterns and genetic associations, supporting a broad, protective effect of crossovers on aneuploidy risk.

Taken together, our findings reveal an overlapping common genetic basis of female meiotic recombination, aneuploidy, and reproductive aging traits that does not measurably intersect with that of clinically diagnosed cases of female infertility. We conclude that common variation in key meiosis genes modulates the high baseline rates of maternal age-associated aneuploidy and thus pregnancy loss within our species.

## Discussion

Pregnancy loss is common in humans ^7^ and often traces to aneuploidy originating in the maternal germline ^1^. Notably, female meiosis initiates in fetal development, when homologous chromosomes pair and establish crossovers, but arrests in this conformation for decades until ovulation and fertilization. Abnormal number or placement of crossovers predisposes oocytes to chromosome mis-segregation upon meiotic resumption ^5,45^. Despite this understanding, the role of common genetic variation in modulating these important molecular processes in humans has remained poorly understood. Through retrospective analysis of large-scale PGT data from human IVF embryos, we simultaneously mapped genetic variants associated with crossover and aneuploidy phenotypes, revealing an overlapping genetic basis involving key meiosis genes.

While we measured significant overdispersion in the age-adjusted rate of aneuploidy per patient and identified significant genome- and transcriptome-wide significant associations, we were intrigued to find that the SNP heritability of aneuploidy was negligible. This finding aligns with low reported SNP heritabilities of female infertility phenotypes ^77^, as well as quantitative genetic theory predicting outsize contributions of environmental and rare genetic variation in fitness-related traits ^78,81^. Nevertheless, given that common and rare variation often converge on the same genes and mechanisms ^82,83^, our results may help inform sequencing-based studies of aneuploidy phenotypes ^48,84,85^. Consistent with mechanistic convergence, rare loss-of-function mutations in several of the genes implicated here have also been linked to meiotic defects and reproductive disorders in smaller clinical cohorts ^86–88^. It is also plausible that a fraction of phenotypic variance for aneuploidy risk could trace to common genetic variation that is inaccessible to genotyping arrays and/or short-read sequencing, for example within technically challenging loci such as large segmental duplications, telomeres, or centromeres. Recent work offered preliminary evidence that particular centromeric haplotypes are enriched among cases of Trisomy 21 ^89^. Future applications of long-read sequencing in PGT may enable validation of this hypothesis and extension to the majority of aneuploidies that are inviable during embryonic development.

The observation that alleles associated with lower rates of recombination are associated with higher rates of aneuploidy raises interesting questions about the evolutionary forces that shape recombination and aneuploidy within and between species. In addition to generating new combinations of alleles, research has demonstrated that recombination is mutagenic, inducing point mutations and structural variation near hotspots of double-strand breaks ^22,90^. These observations together suggest a model of stabilizing selection, whereby rates of recombination may be constrained on the lower and upper ends to limit aneuploidy and other classes of deleterious mutations, respectively. More comprehensive models of recombination rate evolution must also consider the role of crossovers in facilitating adaptation ^91^. By examining patterns of divergence across a mammalian phylogeny, a recent study reported signatures of pervasive positive selection on all meiotic components of the cohesin complex (*SMC1B*, *RAD21L1*, *REC8*, and *STAG3*), which the authors speculated could be explained by intragenomic conflict ^92^. More broadly, the asymmetry of female meiosis is susceptible to meiotic drive, as alleles that enhance their segregation to the oocyte versus the polar bodies will be favored, with examples documented in non-human systems ^93^. The role of meiotic drive in the origins of human aneuploidy remains an important open question.

In summary, our work provides a more complete understanding of common genetic factors that influence risk of aneuploidy—the leading cause of human pregnancy loss. These findings highlight the interplay among the forces of mutation, recombination, and natural selection that operate prior to birth to shape human genetic diversity.

## Data Availability

Aneuploidy and crossover calls are available on Zenodo: https://doi.org/10.5281/zenodo.15114528. Questions regarding clinical genetic testing and raw data should be addressed to Zachary Demko.

## Data and code availability

Genotyping and imputation code is available on GitHub: https://github.com/mccoy-lab/natera_genotyping/. Pipelines for inferring crossover recombination across sibling embryos is available on GitHub: https://github.com/mccoy-lab/natera_recomb. Code for inferring aneuploidies and performing downstream analyses is available on GitHub: https://github.com/mccoy-lab/karyohmm and https://github.com/mccoy-lab/natera_aneuploidy. Aneuploidy and crossover calls are available on Zenodo: https://doi.org/10.5281/zenodo.15114528. Questions regarding clinical genetic testing and raw data should be addressed to Zachary Demko.

## Acknowledgments

Thank you to all individuals who contributed data used in this study. Thank you also to Advanced Research Computing at Hopkins for computing support, as well as Carl Wu, Erik Andersen, Yumi Kim, Yuan He, Dmitri Petrov, Karen Schindler, Jinchuan Xing, and members of the McCoy lab and Origins of Aneuploidy Research Consortium for helpful input. Thank you to George Gemelos and Dusan Kijacic for assistance with data collection. This work is supported by National Science Foundation Graduate Research Fellowship (1746891) to SAC, a Lalor Foundation Postdoctoral Fellowship to AB, a National Institutes of Health (NIH NIGMS) grant R35GM149291 to Carl Wu, the Novo Nordisk Foundation grant NNF22OC0074308 to ERH, a Catalyst Award from Johns Hopkins University to RCM, and a National Institutes of Health (NIH NIGMS) grant R35GM133747 to RCM. The content is solely the responsibility of the authors and does not necessarily represent the official views of the National Institutes of Health.

## Author contributions

RCM initially conceived the project with input from ERH, AB, and SAC. ZPD contributed to collection of the data. Algorithms for inferring aneuploidy and recombination were developed by AB with input from SAC. Parental genotyping and imputation were performed by SAC and AB. Statistical modeling of recombination was performed by RCM and AB. Recombination GWAS was performed by AB. Aneuploidy GWAS, functional genetic analysis of association hits, and phenotype-wide association analyses were performed by SAC. TWAS was performed by MRS. EMSA was performed by XT. SAC, AB, and RCM wrote the paper and generated the figures. All authors read and approved the manuscript.

## Methods

### Data description

#### Ethics statement

The Johns Hopkins University Homewood Institutional Review Board (IRB) determined that this research did not qualify as federally regulated human subjects research and therefore did not require IRB approval (HIRB00011705). This determination was made with the understanding that the research (1) does not involve a systematic research investigation designed to develop or contribute to generalizable knowledge, or (2) does not obtain information or biospecimens through intervention or interaction with a human participant, and use, study, or analyze the information or biospecimens; or does not obtain, uses, study, analyze, or generate identifiable private information or identifiable biospecimens. Data collection and analysis was carried out in compliance with Natera’s IRB approved protocol (Salus #10806) involving Category 4 Exempt Research.

#### Data collection and sampling

IVF samples were collected as part of standard-of-care PGT-A testing, and data was generated using Natera’s CAP-CLIA accredited clinical testing workflow. After fertilization, trophectoderm cells were biopsied from embryos at the blastocyst stage according to the standard protocols of each IVF clinic. Samples were then shipped overnight to the Natera laboratory for PGT-A. Fractions were thawed at 22°C and Arcturus PicoPure Lysis Buffer (Molecular Devices, Sunnyvale, CA, USA) was added to each of the biopsies. The tubes were incubated at 56°C for 1 h and then heat-inactivated at 95°C for 10 min. DNA from the lysed biopsies was amplified using a commercial kit (GE Healthcare, Waukesha, WI, USA) for multiple displacement amplification (MDA). MDA reactions were incubated at 30°C for 2.5 h and then heat-inactivated at 65°C for 5 min. The amplified samples were genotyped using Illumina (San Diego, CA, USA) Infinium II genotyping microarrays (CytoSNP-12 chips) using a modified 24-h protocol, as described previously 115. Parent buccal samples were collected using MasterAmp Buccal Swabs (Madison, WI, USA). Genomic DNA was isolated from these swabs using Epicentre DNA Extraction Solution (Madison, WI, USA). For parental samples, the standard Infinium II protocol (www.illumina.com) was used. Patient level genetic data and summary metadata (including parental ages, egg and sperm donor statuses, and year of sample collection) were de-identified and provided by Natera to Johns Hopkins for analysis.

#### Sample overview

After initial quality control (presence of files in dataset, removing families with listed ages of biological parents outside of 18-90 years old), the dataset included 22,850 unique biological mothers with data collected from 2014-2020 (between 2,271 and 4,719 unique mothers per year). Maternal ages ranged from 20.1 to 55.8 years at the time of collection. Excluding cases that used egg donors, the mean maternal age was 36.2 years (**Fig. S1**). Each IVF cycle had a mean of 4.63 embryos (standard deviation = 3.40; range = 1-35). Most pairs of biological parents (17,420) had one recorded cycle, 4,021 had two cycles, and 1,409 had three or more.

### Genotyping, imputation, and quality control

We restricted our analysis to samples with genotype data recorded for all array probes. Starting with raw array probe intensity values (x,y), we applied the recommended Illumina normalization procedure:

1. Exclude outliers by being outside the 99th and 1st quantiles of the distribution of x, y, or x/(x+y) across all probes on a chromosome
2. Correct for x and y offsetting from (0,0)
3. Correct for rotational angle from the x-axis (theta)
4. Correct for rotational angle from the y-axis (shear)
5. Scale x,y points by axis-specific mean estimates

The normalization procedure ensures that all intensities are on the same approximate scale prior to genotyping. Following this procedure, we also filtered out ultra-rare variants with global allele frequencies less than 0.1% (15,534 variants removed).

We then used the program optiCall ^95^ to call genotypes from the normalized array intensities. To avoid population structure driving deviation from Hardy-Weinberg equilibrium, we provided super-population labels derived from k-means clustering of PCA on the raw intensity values (*K*=3) when calling genotypes. We split each chromosome into segments with approximately 500 genotyped SNPs per chunk. We restricted output to genotypes with posterior probabilities of at least 0.9 (-minp 0.9) and used Hardy-Weinberg equilibrium p-values greater than or equal to the default threshold of 1 × 10^-15^. After calling genotypes, we lifted over variants from human genome build GRCh37 to GRCh38, which resulted in the removal of 1,831 variants. Following application of these filters, we retained 275,425 variants across the genome.

In preparation for genotype imputation, we pre-phased the parental genotypes using Eagle v2.4.1 ^96^ and the combined Human Genome Diversity Project and 1000 Genomes Project (HGDP + TGP) reference panel haplotypes ^97^. We then applied genotype imputation with BEAGLE v5.4 ^98,99^ using the same HGDP + TGP reference panel with default parameters. Each autosome was split into 20 non-overlapping intervals for speed. For the X chromosome, we split the samples into 10 equal groups for more efficient memory management when running imputation. After imputation, we retained variants with dosage r-squared value greater than 0.8 ^100^.

To evaluate the broad ancestry composition of the parental samples, we combined these genotype data with published data from 2,504 unrelated individuals from the 1000 Genomes Project, restricting to an overlapping set of 257,580 autosomal biallelic variants. We then performed principal component analysis on the combined genotype matrix (**Fig. S2A**). For the purpose of **Fig. S2B**, samples were labeled based on genetic similarity, using the majority of ancestry labels of the 5 nearest neighbors based on Euclidean distance across the top 20 principal components, scaled by the percentage of variance explained, to reference individuals from the 1000 Genomes Project.

### Aneuploidy and crossover detection

#### A haplotype-copying HMM for aneuploidy detection from allelic intensity data

To model the relationship between array intensities, parental haplotypes, and the underlying ploidy of an embryo chromosome, we formulated a hidden Markov Model (HMM). The hidden states *Z*_*i*_ are tuples representing the maternal *M* = *M* = (*m*^(0)^, *m*^(1)^) and paternal *P* = (*p*^(0)^, *p*^(1)^) haplotypes that are copied at the *i*^*th*^ locus. We detail all the possible hidden states per karyotype class below:

- Nullisomy: ⊘
- Maternal Monosomy: {*p*^(0)^, *p*^(1)^}
- Paternal Monosomy: {*m*^(0)^, *m*^(1)^}
- Disomy: { (*m*^(0)^, *p*^(0)^), (*m*^(0)^, *p*^(1)^), (*m*^(1)^, *p*^(0)^), (*m*^(1)^, *p*^(1)^) }
- Maternal Trisomy: { (*m*^(0)^, *m*^(0)^, *p*^(0)^), (*m*^(0)^, *m*^(1)^, *p*^(0)^), (*m*^(1)^, *m*^(1)^, *p*^(0)^), (*m*^(0)^, *m*^(0)^, *p*^(1)^),(*m*^(0)^, *m*^(1)^, *p*^(1)^), (*m*^(1)^, *m*^(1)^, *p*^(1)^) }
- Paternal Trisomy: { (*m*^(0)^, *p*^(0)^, *p*^(0)^), (*m*^(0)^, *p*^(1)^, *p*^(0)^), (*m*^(0)^, *p*^(1)^, *p*^(1)^), (*m*^(1)^, *p*^(0)^, *p*^(0)^),(*m*^(1),^ *p*^(0)^, *p*^(1)^), (*m*^(1)^, *p*^(1)^, *p*^(1)^) }

For example, for a maternal monosomy at locus *i*, the model can only copy from either *paternal* haplotype *p*_*i*_^(0)^ or *p*_*i*_^(1)^, since the maternal chromosome is absent. The variable *M*_*i*_ is the allelic dosage (i.e., the number of alternative alleles) of maternal-origin haplotypes copied at locus *i*; this is analogously defined for the paternal dosage *P*_*i*_. The variable *K*_*i*_ is the total ploidy (i.e., the size of the tuple defining *Z*_*i*_). Using these auxiliary variables, we define the emission distribution for the observed allelic intensity at the *i*^*th*^ locus, *b*_*i*_:

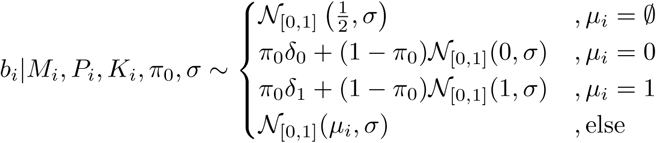

where 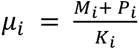 is the expected dosage *conditional* on the parental haplotypes being copied (the case of *μ*_*i*_ = *∅* for a nullisomy). Since the range of allelic intensity is between 0 and 1, we model the emission using a truncated normal distribution (*N*_[0,1]_) or mixture of a point-mass and a truncated normal distribution. The technical noise parameters *π*_0_ and *σ* reflect 1) the fraction of fully homozygous genotypes lying at the allelic intensity boundaries of 0 or 1 and 2) the standard deviation in the intermediate allelic ratios, respectively.

To complete the HMM definition, we define the transition matrix *A*. There are two different classes of transitions between hidden states: 1) transitions *within* the same ploidy class and 2) transitions *between* ploidy classes. In the first case, this is likely due to a recombination event, which occurs with rate *r*. In the second case, inter-ploidy class transitions occur with probability *a*. We assume throughout that *r* >> *a* and set *r* = 10^−8^ per base pair per generation as an estimate of the genome-wide recombination rate (given that 1 centimorgan ≈ 1 Mbp in humans) and *a* = 10^−10 22^. This means that an inter-ploidy transition is ∼100 times less likely than recombination between loci and requires strong evidence over a longer stretch of the chromosome. Using these two variables, *A* is defined as:

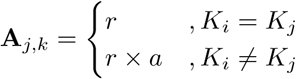

where *K*_j_ is the ploidy count of latent state *j* and *A*_*i*,*i*_ = 1 − ∑_j≠*i*_ *A*_*i,j*_. The full transition probability is therefore:

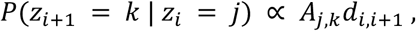

where *d*_*i*,*i*+1_ is the physical distance in base pairs between locus *i* and *i* + 1. Using the forward algorithm to compute the likelihood of the data *b̄*, we obtain maximum-likelihood estimates 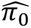 and 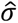 using the bounded BFGS algorithm for numerical optimization ^101^. Using the MLE parameters 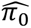 and 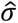, we calculate the posterior probability of being in each state 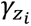 at locus *i* via the forward-backward algorithm ^101^. We calculated the posterior probability of a given ploidy context as the scaled posterior probability of ploidy across all sites on the chromosome. For the case of a disomy:

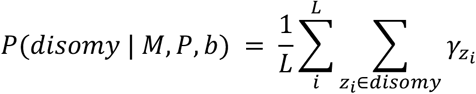

The posterior probability of the other ploidy classes is similarly calculated. Following the calculation of the posterior probability of each ploidy configuration, we assign the ploidy configuration with the maximum posterior probability as the karyotype for the chromosome in that embryo. Downstream, we use the maximum posterior probability > 0.9 as a threshold for high-confidence whole-chromosome aneuploidy.

#### Sex chromosome aneuploidy detection

To accommodate calling sex chromosome aneuploidies, we modify the hidden states of the HMM for the X chromosome:

- Loss of X: ⊘
- Single paternal copy of X: {*p*^(0)^}
- Single maternal copy of X: {*m*^(0)^, *m*^(1)^}
- Disomic Bi-parental X chromosome inheritance: { (*m*^(0)^, *p*^(0)^), (*m*^(1)^, *p*^(0)^)}
- Uniparental Disomy of maternal X chromosome: { (*m*^(0)^, *m*^(0)^), (*m*^(0)^, *m*^(1)^), (*m*^(1)^, *m*^(1)^)}
- Trisomy of X chromosome: { (*m*^(0)^, *m*^(0)^, *p*^(0)^), (*m*^(0)^, *m*^(1)^, *p*^(0)^), (*m*^(1)^, *m*^(1)^, *p*^(0)^)}

To adjust the model for the Y chromosome we only use two hidden states:

- Loss of Y: ⊘
- Presence of Y: {*p*^(0)^}

We used the same transition model as was used for the autosomes: a recombination rate of 10^−8^ per base pair per generation and inter-ploidy transition rate of 10^−10^. Posterior probabilities of chromosome-wide ploidy states are obtained by collapsing the results of the forward-backward algorithm. Karyotype status is assigned as the maximum posterior probability, and we similarly filter on posterior probability > 0.9 to obtain high-confidence whole-chromosome aneuploidy calls.

#### Performance evaluation of aneuploidy detection

To evaluate the performance of our method for estimating karyotypes, we simulated array intensity data assuming various whole-chromosome ploidy states. Specifically, we simulated parental haplotypes with 4,000 SNPs along a contig of 35 Mbp (a close approximation to the real data on chromosome 22 for parental haplotypes). To reflect ascertainment of alleles in array data, we drew parental alleles under Hardy-Weinberg equilibrium from the distribution of global allele frequencies of variants on the Illumina HumanCytoSNP array from the 1000 Genomes Project ^102^. To account for switch error rates consistent with population-based phasing, we simulated a switch-error rate of 3% across parental haplotypes, representing an upper-bound of expected phasing errors ^103^. Under the parameters *π*_0_, *σ*, we simulated 40 replicates each of nullisomy, monosomy, disomy, and trisomy. As the copying probability is symmetric across the sexes, we only focus on simulating monosomy and trisomy of paternal origin for performance evaluation.

Under the case of true meiotic aneuploidies, across the entire range of parameters the precision and recall of the current method are > 0.95 across all categories simulated, indicating high accuracy for detection of aneuploidies while accounting for genotyping array-specific noise (**Fig. S3**).

Early embryos may also be affected by mosaic aneuploidies, where only a fraction of the cells in the biopsy are aneuploid and others are disomic ^104,105^. To assess the potential confounding impacts of mosaic aneuploidies, we simulated both 5 and 10-cell biopsies, where the expected proportion of cells containing a specific aneuploidy is *p*_*mosaic*_. We simulated 20 replicates across *p*_*mosaic*_ from 0% to 100% of the cells containing an aneuploid set of chromosomes. Each mosaic cell that is simulated represents a single focal aneuploidy (either monosomy or trisomy). We then averaged the allelic intensities across all the cells (disomic and non-disomic) to create the vector of allelic intensities for inference.

We find that simulated mosaic aneuploidies tend to exhibit different effects depending on whether the mosaic aneuploidy is a monosomy or trisomy and their fraction in the dataset. For mosaic trisomies, once the cell fraction increases to approximately 50%, we observed confident trisomy calls along the whole chromosome irrespective of genotyping array noise (**Fig. S15**). Monosomies behave slightly differently, where mosaic monosomies with a cell fraction between 10% and 80% tend to be confidently called as trisomies (**Fig. S15**). This is because a mixture of monosomic and disomic chromosomes create modes in the distribution of allelic intensities that are similar to those of a true meiotic trisomy (e.g., peaks in allelic intensity ratios at ⅓ or ⅔). When the cell fraction of monosomy is > 80%, we find that a monosomy is confidently called with a posterior probability > 0.9. Therefore, while our method performs well for detection of meiotic aneuploidies, certain forms of mosaic aneuploidy may impact performance, motivating additional filters.

#### Filtering mosaic aneuploidies

To statistically separate mosaic (mitotic-origin) from meiotic aneuploidies, we exploit two well-characterized signatures of mitotic aneuploidies. The first signature is that mitotic trisomies only display *single parental homologs* (SPH) from a given parent, whereas meiotic-origin trisomies contain *both parental homologs* (BPH) from one of the parents and thus three genetically distinct parental haplotypes ^8,20,106^. The second signature is that mitotic aneuploidies exhibit no strong relationship with maternal age ^37^. For trisomic calls made under the HMM model, we estimate the posterior probability of a chromosome being in a BPH vs. SPH state along the entire length of the chromosome. It should be noted that the BPH vs. SPH is not a perfect indicator of meiotic vs. mitotic trisomies (or mosaic monosomies), as tracts of BPH may exist in distal regions of chromosomes, outside of the range of the array ^106^. Similarly, meiosis II errors with no recombination may manifest as SPH trisomies. In order to distinguish mitotic from meiotic aneuploidies, we investigated at what level of BPH the maternal age effect becomes significant.

For each confidently called trisomy (posterior probability > 0.9), we calculated the posterior probability of BPH and considered every trisomy call with *P*(*BPH* | *Data*, *Θ*) < *α* a mitotic-origin aneuploidy. We then ran a binomial regression for the number of inferred mitotic aneuploid embryos against maternal age and tested at what level *α* the effect-size for the linear effect becomes significantly positive (**Fig. S16**). We tested across a grid of 100 points for *α* from 0 to 1 to determine when 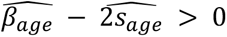 which are the estimated effect size and standard error for the age effect, respectively. We expect at higher thresholds of BPH that meiotic aneuploidies will begin to be included in this filter, making the age effect non-zero. We find that a posterior filter of *P*(*BPH* | *Data*, *Θ*) < 0.340 maintains the null effect of maternal age for inferred mitotic aneuploidies (**Fig. S16**). This filter likely captures mitotic-origin trisomies affecting all cells of a given biopsy, as well as mosaic trisomies and monosomies of medium cell fractions.

#### Segmental aneuploidy detection and filtering

To prioritize aneuploidies affecting whole chromosomes, we developed a pipeline to identify sub-chromosomal (i.e., "segmental") aneuploidies using the HMM output and exclude these chromosomes from downstream analyses. Using the maximum *a posteriori* (MAP) path through the HMM, we identify changepoints in the path that support different karyotypic states (i.e., inter-ploidy transitions). Smaller segmental aneuploidies may potentially be present on longer chromosomes but may not represent enough of the MAP path across the chromosome to shift the total posterior across all sites to be < 0.9, which we use to assign whole-chromosome aneuploidy status.

We identify changes in the MAP path from the majority ploidy of the chromosome and determine whether it is a segmental aneuploidy based on 1) whether it contains at least 100 SNPs, 2) whether the local posterior within the segment supporting its ploidy assignment is > 0.8 across the segment, and 3) whether the segment is at least 5 Mbp long. We find that this set of filtering criteria has reasonable power for identifying simulated segmental aneuploidies that are > 5 Mbp in length with a precision > 90% across different scales of embryo-level noise (**Fig. S17**). We exclude all chromosomes with segmental aneuploidies called in this way from our current analyses of whole-chromosome meiotic aneuploidy. Conceptually, many changepoints occurring in the MAP path indicate either 1) biopsy errors or 2) potential mosaic aneuploidies. In either case, we expect that both of these occurrences will prevent an aneuploidy from reaching a chromosome-wide posterior probability > 0.9.

#### Calling crossover recombination events in PGT-A data

To identify crossover events, we used a previously defined heuristic approach, which uses switches in the assignments of informative variants from each parent to designate the endpoints of a crossover in a template embryo relative to its sibling embryos ^107^. For each chromosome, we restrict our analysis to only families that have ≥ 3 disomic embryos for that chromosome (noting that embryos may have aneuploidies occurring on other chromosomes).

Previous methods relied on *called genotypes* for siblings, which are not reliably available for PGT-A data. Therefore, we adapted the method to consider the likelihood that the same parental allele is transmitted to the non-template sibling embryo. Our approach is based on the likelihood for the embryo genotype array intensity, conditional on the parental genotypes (*P*_*i*_, *M*_*i*_ for paternal and maternal, respectively) at site *i*: *L*(*b*_*i*_ | *M*_*i*_, *P*_*i*_, *π*_0_, *σ*). In this case we also use the MLE estimates of 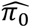, 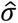 for the embryo under consideration. To illustrate, consider two informative biallelic SNPs (*a*, *b*) for a maternal crossover event—where the maternal genotypes are heterozygous and the paternal genotypes are homozygous—at which we can compute the following likelihood of the *same allele being transmitted* to both the template (*t*) and a single non-template embryo (*nt*):

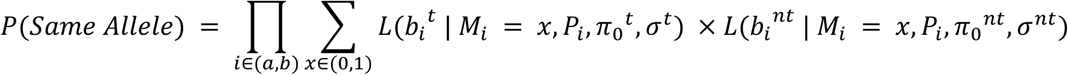

The likelihood for *different alleles being transmitted* from the maternal side at (*a*, *b*) for the template and non-template embryos is:

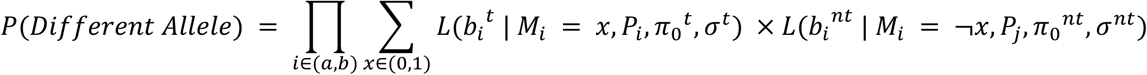

Thus, for every pair of *t*, *nt* embryos within the family, we can determine whether a crossover event occurred in the transmission from the mother to the template embryo between *a* and *b* by evaluating the log-likelihood ratio *llr*_{*a*,*b*}_ = *log*(*P*(*Same Allele*)) − *log*(*P*(*Different Allele*)). Following previous work, we create a binary set of “switch indicators’’ of *llr*_{*a*,*b*}_ < 0 or *llr*_{*a*,*b*}_ > 0 between each pair of informative markers to isolate switch points indicative of crossover events ^107^.

To leverage multiple sibling embryos and account for noise on the genotyping array, we apply two further refinement steps to isolate crossovers. The first step considers up to five adjacent pairs of informative sites and determines whether the number of switches within the switch cluster is odd, since an even number of switches (very close and nearby crossover) can be indicative of genotyping error ^107^. The second filter restricts crossovers to be supported by the majority of the sibling embryos to ensure that multiple meioses from the parental individual support a crossover call.

### Genome-wide and transcriptome-wide association with recombination phenotypes

Recombination phenotypes were defined in the following categories on a sex-specific and joint basis:

1. <u>Recombination Rate</u> For each meiosis from a given parent, we estimated the total number of crossovers across all of the autosomes. We define the phenotype as the mean number of autosomal crossovers across all euploid embryos.
2. <u>Hotspot Occupancy</u> We first defined hotspots as regions where the sex-averaged genetic map has a relative recombination rate > 10, following previous definitions ^22^. We then estimate the *fraction* of crossover intervals that intersect with hotspots after permuting crossover intervals ^107^. The phenotype is an approximation to the maximum likelihood estimate of the fraction of crossovers of a specific parental origin (across all meioses observable for that parent) that occur in hotspots.
3. <u>Replication Timing</u> Replication timing was derived from measurements in 300 induced pluripotent stem cell (iPSC) lines ^108^. Coordinates were lifted over to GRCh38. Using the average replication timing across all 300 iPSCs, we used linear interpolation to estimate the replication timing at each crossover interval midpoint. Replication timing for an individual was estimated as the mean replication timing value across all corresponding crossover intervals.
4. <u>GC Content</u> Guanine-cytosine (GC) content at each estimated crossover location was calculated using the GC percentage track in five base pair increments from the UCSC browser in GRCh38 coordinates. We estimated the GC content at each crossover location using the average GC content within 500 bp upstream and downstream of the midpoint of the crossover interval. GC content for each individual was estimated as the average GC content across all crossovers attributed to that individual, across all corresponding meioses.

All phenotypes were inverse-rank normal transformed within each sex-specific study and jointly and tested using REGENIE ^109^. All association testing included age, number of sibling embryos assayed, the average estimated *π*_0_ and *σ* parameters corresponding to the embryos from a given parental pair, and 20 genetic principal components across all the parental individuals as covariates. For joint testing of maternal and paternal phenotypes, we also included sex as a covariate.

We conducted linkage disequilibrium (LD) clumping (plink2 --clump-r2 0.1 --clump-kb 1000) of association results to identify a set of approximately independent variants per phenotype. To map the lead variant within each locus to a gene, we assigned the closest gene and reported the distance to the gene boundary in GENCODE v37 (**Table S2**). To evaluate potential novelty of association results, we evaluated replication at two scales: 1) whether a specific lead variant in a locus replicates, and 2) whether a specific gene is also found to replicate in a previous GWAS of recombination phenotypes ^22^.

We performed GWAS across these 4 recombination phenotypes, stratifying by parental origin as well as considering both parents together (12 total GWAS). We identified 42 approximately independent loci (r^2^ ≤ 0.1) exceeding the threshold of genome-wide significance (*p* < 5 × 10^-8^), implicating 15 unique genes, all of which replicate previous findings in the literature ^22^ (**Table S3**).

TWAS for each recombination phenotype was conducted similarly to the TWAS for aneuploidy (see Methods). We applied a linear model using the glm function (family=gaussian) within R (version 4.3.3^110^) to test the association between the predicted expression levels of genes and each recombination phenotype on a sex-specific and joint basis. Covariates in the model were the same as those used in the GWAS. A combined TWAS p-value was computed for each protein-coding gene across all tissues using the ACAT ^111,112^ method followed by Bonferroni correction for multiple testing (*p* = 0.05 / 16,685 unique protein-coding genes = 3 × 10^-6^).

### Genome-wide and transcriptome-wide association with maternal meiotic aneuploidy

#### Filtering copy number calls

The aneuploidy phenotype is based on karyoHMM (see Methods), which outputs a posterior probability for each copy number state for each chromosome. We only consider chromosomes that have a posterior probability > 0.9. Any chromosome that does not reach that threshold is excluded from the analysis, though remaining chromosomes from the same embryo are still considered. Embryos with a nullisomy call for 5 or more chromosomes were deemed to have inadequate or missing data and are excluded from analysis. We additionally required a minimum Bayes factor of 2, indicating that a given call has at least twice the support of the next most likely copy number state.

The karyoHMM method also estimates the standard deviation of the B-allele frequency (BAF) distribution conditional on parental genotypes, *σ*F, which quantifies the experimental noise in PGT biopsy data (see Methods). To reduce potential experimental noise, we excluded embryos with an average *σ*F outside 3 standard deviations from the cohort-wide mean. The karyoHMM method also identifies segmental aneuploidies (copy number gains/losses only affecting a portion of a chromosome). To ensure our phenotype was focused on whole-chromosome gains and losses, we removed any chromosome that was affected by a segmental aneuploidy, defined as a stretch greater than 5 Mbp with more than 100 SNPs and a greater than 80% posterior probability for a copy number other than disomy (see Methods).

#### Genome-wide association study

To select a set of unrelated individuals for association testing, we used KING ^113^ to exclude all first or second degree relatives. 5,430 female individuals were present in the dataset multiple times, indicating multiple IVF cycles. For the purpose of defining discovery and test sets, we calculated a weighted mean age and total embryo count by merging families that underwent multiple cycles of IVF. After removing duplicate individuals, we randomly assigned 85% of the mothers to the discovery set, verifying that the distributions of maternal age and number of day-5 embryos were similar between the discovery and test sets. This procedure resulted in the assignment of 19,529 unique mothers to the discovery set and 3,447 unique mothers to the test set. We then propagated these assignments to the corresponding partners.

The aneuploidy phenotype was defined on a per-cycle basis. Cycles were inferred based on metadata provided by Natera, in which each unique set of biological parental ages was interpreted to define one IVF cycle. Most often, a single casefile identifying number (casefileID) comprised a single IVF visit. In situations where ages were provided for the mother but not the father or embryos, the ages listed for a mother were propagated to other samples within the same casefileID. For cases that used an egg or sperm donor, the provided parental ages were those of the individuals undergoing IVF rather than the individual providing the egg or sperm. Therefore, egg donors were assigned the average age of egg donors, 25 years, based on a published retrospective study of egg donors spanning years overlapping our study ^114^. Sperm donors were similarly assigned the average age of sperm donors, 27 years, based on a published study ^115^.

An embryo was categorized as aneuploid if it possessed between 1 and 5 autosomes assigned as maternal-origin aneuploid by karyoHMM. To conduct the association test for aneuploidy, we applied a generalized linear mixed-effect model (implemented via glmer function from the R package lme4, version 1.1.35.5 ^116^), where patient ID was included as a random effect grouping factor to account for the fact that 23.76% of female individuals had multiple IVF cycles. We used a binomial family, as the phenotype is encoded as the counts of aneuploid and euploid embryos per cycle. We used the first 20 genotype principal components (PCs), maternal age, paternal age, egg donor status, and sperm donor status as fixed effect covariates. To obtain the PCs, we applied a principal components analysis (PCA) using PLINK (version 1.9) to the parental genotypes (output from optiCall) for all autosomes.

To test whether the incidence of female meiotic aneuploidy is individual-specific, we fit a quasi-binomial generalized linear regression model to the counts of embryos affected versus unaffected with maternal meiotic-origin aneuploidy (combining across cycles per set of biological parents), including a quadratic term to model the relationship with maternal age.

Three female individuals with maternal age greater than 50 years were excluded from this analysis, as they were observed as high-leverage outliers based on Cook’s distance (mean *D* = 0.22). We then simulated new counts of affected and unaffected embryos from the fitted model for the same sample (n = 1,000 simulations), but assuming no overdispersion (i.e., dispersion parameter (φ) = 0). Dispersions were calculated as the sum of squared Pearson residuals, divided by the residual degrees of freedom.

We used the LDproxy tool from LDlink^117^ to evaluate LD between GWAS lead SNPs and other potential causal variants within the genomic region. Specifically, we computed computed pairwise LD between a given query variant and other variants in a ±500 kbp window using the genotype data from high-coverage sequencing of European population samples from the 1000 Genomes Project^118^, aligned to reference genome build GRCh38.

#### Transcriptome-wide association studies (TWAS) of maternal meiotic aneuploidy

We performed TWAS by using the imputed parental genotype data (see Methods) to predict genetically regulated gene expression across each of 49 tissues. The published 49 tissue-specific multivariate adaptive shrinkage in R (MASHR) prediction models were trained on GTEx v8 expression data ^119,120^.

For each gene, we then applied a linear mixed-effects model as described for GWAS above with the same covariates, but here evaluating the association between counts of aneuploid versus euploid embryos and predicted gene expression. We repeated this procedure for each tissue, then combined the single-tissue TWAS p-values using the Aggregated Cauchy Association Test (ACAT) ^121^. Multiple hypothesis testing correction was performed using a Bonferroni correction for the number of protein-coding genes across all tissues (*p* = 0.05 / 16,685 unique protein-coding genes = 3 × 10^-06^).

We investigated the extent to which co-regulation might lead to multiple association signals at a TWAS locus by plotting the expression of pairs of genes across individuals from published GTEx v8 expression data ^41^.

### Quantifying nucleus-wide covariation in crossovers between euploid and aneuploid embryos

To establish evidence of per-nucleus covariation in crossover counts per embryo, we used simulation and variance decomposition routines from previous research on individual gametes ^44^. To compare against independent simulations, for each of the 46,861 euploid embryos with estimated crossovers, we created an independent crossover count by drawing each autosomal crossover count from the full pool of embryos assayed for that chromosome, restricted to the appropriate parent. We observe that for both maternal and paternal crossovers, there is substantial overdispersion in crossover counts, consistent with previous findings in oocytes ^44^ (**Fig. S9**).

To decompose the per-embryo variance in crossovers, we turn to a decomposition of the variance in crossover number, where the total variance (*A*) is decomposed into the independent component of variance on a chromosome (*B*) and the covariance *between* chromosomes in crossover count (*C*) :

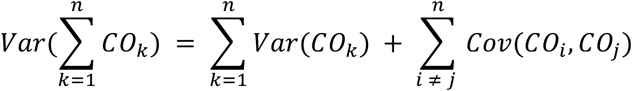

A ratio of *C/A* >> 0, which we term inter-chromosomal covariance (ICC), implies that a substantial fraction of the variance in autosome-wide crossover counts is contributed by positive covariance between crossovers on individual chromosomes.

| | $A$ | $B$ | $C$ | $C/A$ |
| --- | --- | --- | --- | --- |
| Maternal | 154.520 | 50.778 | 103.741 | 0.671 |
| Paternal | 105.185 | 35.861 | 69.324 | 0.659 |

To compare with classical statistical estimators, we also computed the intra-class correlation (also ICC) in crossover counts stratified by parental origin, where the classes are defined as the per-chromosome crossover counts and are grouped by individual embryo identifiers. The intra-class correlation in this case is significantly non-zero for both maternal (0.176; *p* < 10^-100^) and paternal crossovers (0.088; *p* < 10^-100^).

### Mixed-effect models to contrast crossovers between euploid and aneuploid embryos

To compare the crossover counts between euploid and aneuploid embryos on their corresponding disomic chromosomes, we used mixed-effect models that include nested random effects for both the parental individual and the embryo in question. For crossover counts, we use a Poisson mixed-effect model with a log link function:

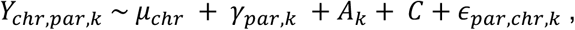

where *Y*_*chr*,*par*,*k*_ is the crossover count on a specific chromosome *chr* for parent *par* and embryo index *k*. The random effect G_*par*,*k*_ is a nested random effect for embryo-specific variance nested within parent-specific variance components in crossover counts (i.e., (1 | par / k) in R). Fixed effects included the expected rate of crossovers per chromosome (*μ*_*chr*_) using the total centimorgan distance as a fixed covariate, an indicator of whether the embryo contained an inferred maternal meiotic aneuploidy (*A*_*k*_), and female individual-specific covariates (*C*) which include ancestry principal components, maternal age, and average estimates of embryo noise parameters from karyoHMM.

The model was fit using REML in the glmer package in R (version 4.4.1). For reporting results, we use the estimated marginal means (via the emmeans package) for the binary indicator *A*_*k*_ of an inferred maternal meiotic aneuploidy affecting the *k*^*th*^ embryo.

To assess the effect of patient-specific crossover rates on aneuploid embryos, we first extracted the estimated mean parental random effects (across both euploid and aneuploid embryos) from the above Poisson mixed-effect model 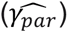. We then used the per-patient crossover rates in a binomial mixed-effect model to test whether donor-specific crossover rates were associated with the proportion of aneuploid embryos, while adjusting for principal components, parental ages, parental age squared, donor status, and the genotype for each of the significant SNPs included in our phenome-wide association study.

### Heritability estimation for recombination and aneuploidy phenotypes

The SNP heritability for maternal meiotic aneuploidy and recombination phenotypes was estimated using LD-score regression with LD scores computed from our sample genotype data ^122^. SNP-heritability for age at menopause and age at menarche was estimated using LD-score regression using LD scores computed on individuals assigned to the EUR continental group within the HGDP + TGP reference panel ^97^ as well as summary statistics from the ReproGen consortium ^70,71^. SNP heritability estimates for body mass index and height were downloaded from the pan-UK Biobank project ^123^. Heritability estimates for infertility traits were accessed from published data ^77^. Clinical definitions of infertility followed those from Venkatesh et al. (see their Supplement 3, Table 1 for clinical criteria) ^77^.

### Electrophoretic mobility shift assay

Using MAST from the MEME suite ^60^, we found that the reference genome sequence surrounding a fine-mapped eQTL of *SMC1B* (rs2272804; **Fig. 3D**) is a predicted binding motif for the transcription factor ATF1, whereas the sequence including the alternative allele is not. To test this potential reduced binding activity *in vitro*, we conducted an electrophoretic mobility shift assay (EMSA) with four DNA sequences: the sequence of *SMC1B* surrounding the putative ATF1 binding site and containing the REF allele of rs2272804, the same sequence but substituting the ALT allele of rs2272804, an ATF1 consensus binding sequence (positive control ^124^), and a sequence with no predicted ATF1 binding activity (negative control).

The forward sequence of the DNA fragments are recorded here, with the ATF1 motif bolded and underscored. The variant base is the last position of this motif (C → A).

- Sequence centered on rs2272804 with REF allele (30 bp)

- 5′-TGTACCTCTGCGG**<u>CGTCAC</u>**TGGGAGCCCGA-3′
- Sequence centered on rs2272804 with ALT allele (C → A) (30 bp)

- 5′-TGTACCTCTGCGG**<u>CGTCAA</u>**TGGGAGCCCGA-3′
- Positive control: ATF/CRE consensus (30 bp) from the Epstein-Barr virus LMP1 gene promoter

- 5′-TCTAGCTCTCTGA**<u>CGTCAG</u>**GCAATCTCTGA-3′
- Negative control: HSPA1A (hsp70) promoter (35 bp)

- 5′-ATCGAGCTCGGTGATTGGCTCAGAAGGGAAAAGGC-3′

DNA oligonucleotides were ordered from IDT for both the forward and reverse strand, with the forward oligonucleotides labeled at the 5’ with FAM. Oligonucleotides were dissolved in duplex buffer (100 mM potassium acetate; 30 mM HEPES, pH 7.5) and complementary oligonucleotides were annealed to double-strand fragments in a thermocycler by heating up to 95°C for 5 min, then ramping down to 20°C at 5°C/min.

EMSA was used to determine the *K_D_* of ATF1 to DNA fragments. 5nM DNA were mixed with various concentrations of recombinant ATF1 protein (SinoBiological, A09-54G) in the presence of 50 ng poly(dI-dC) (Thermo Fisher Scientific, 20148E) in a 10 µl reaction containing 25 mM HEPES pH 7.5, 50 mM KCl, 50 mM NaCl, 5 mM MgCl2, 5% glycerol, 1 mM DTT, 0.01% IGEPAL, 0.25mM TCEP, and 250 ng/µl BSA. The reactions were incubated at 37°C for 30 min, then analyzed by electrophoresis on 2% agarose gel run in 0.2x TB buffer (17.8 mM Tris and 17.8 mM boric acid) at 100V for 45 min. Gels were scanned on a Typhoon 5 Variable Mode Imager (GE Biosciences), and bands were quantified using the Image Studio software (LICORbio) to calculate *F_bound_* (fraction of DNA bound) as *I_bound_*/(*I_bound_* + *I*_un_*_bound_*), where *I* is the intensity of the corresponding band.

The *F_bound_* in each reaction was plotted versus the concentration of protein, and the data were fit with the binding equation *F_bound_* = *B_max_*([ATF1]/(*K_D_* + [ATF1])) by non-linear regression to estimate the value of *K_D_*, where *B*_max_ (the fraction bound at which the data plateaus) was assumed to be 1 ^125^.

