## Supplementary Figures for "Common variation in meiosis genes shapes human recombination phenotypes and aneuploidy risk"

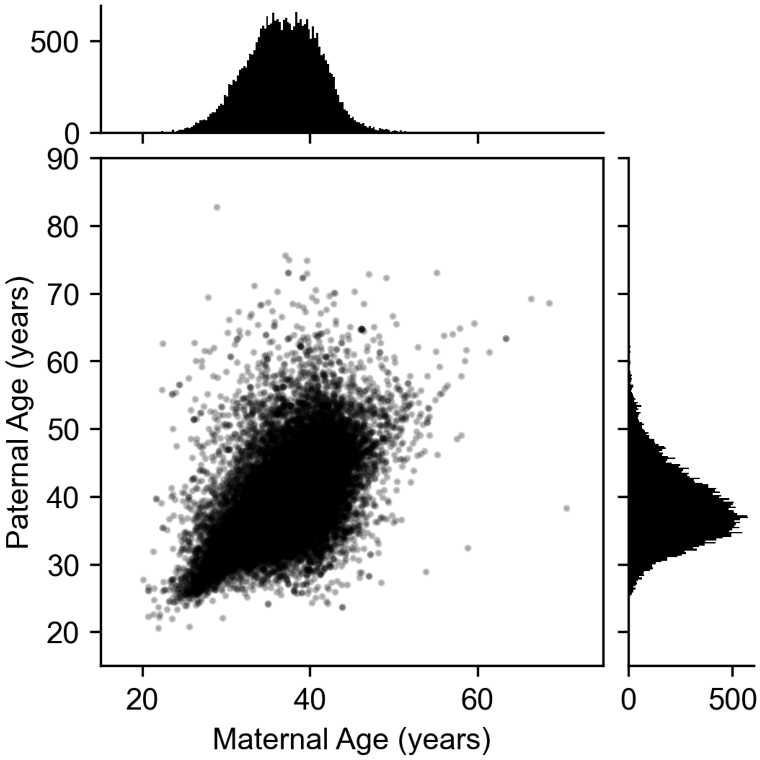


**Figure S1**. Age at time of collection across 22,850 sets of biological parents.

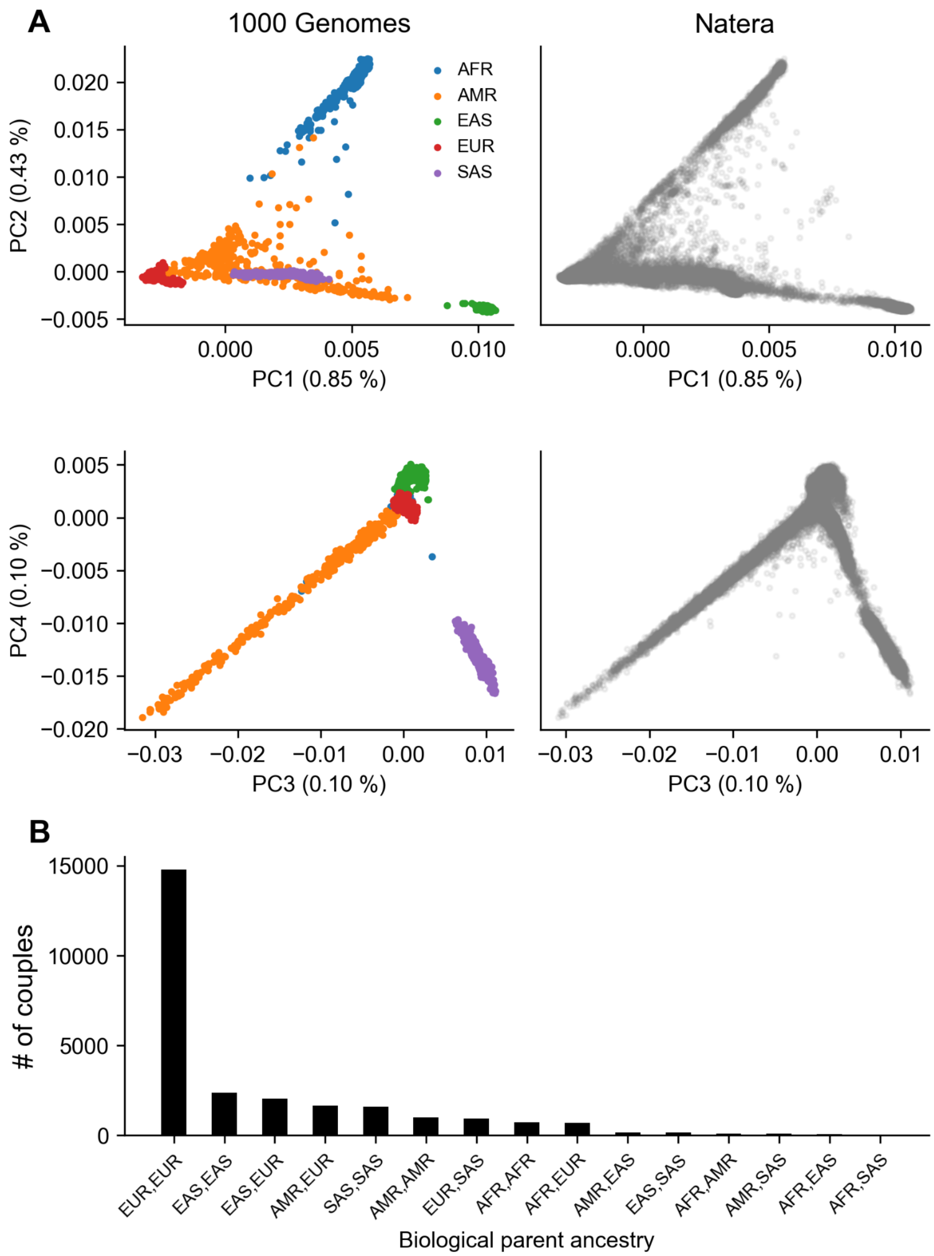


**Figure S2**. Population structure and ancestry inference of biological parents in the Natera datasets. (A) Joint PCA with 1000 Genomes individuals at shared polymorphic sites on the HumanCytoSNP array for only participants and partners, excluding embryos where genotypes cannot be reliably assigned. (B) Ancestry of biological parents, where labels reflect increased genetic similarity to reference individual population labels from the 1000 Genomes Project regional groupings using nearest-neighbors in PCA-space.

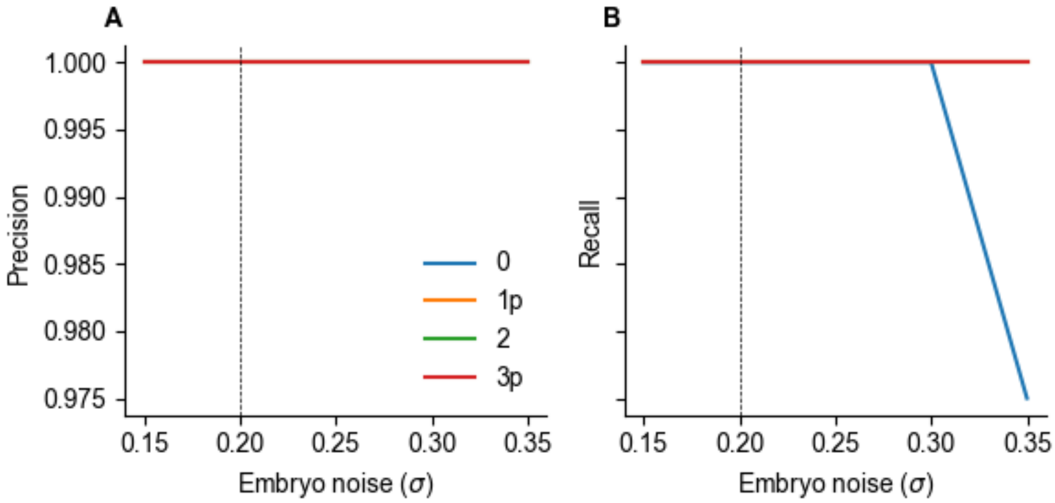


**Figure S3.** Performance of the karyoHMM method to detect simulated meiotic aneuploidies. Dashed vertical line represents the median maximum-likelihood noise parameter across all real embryos.

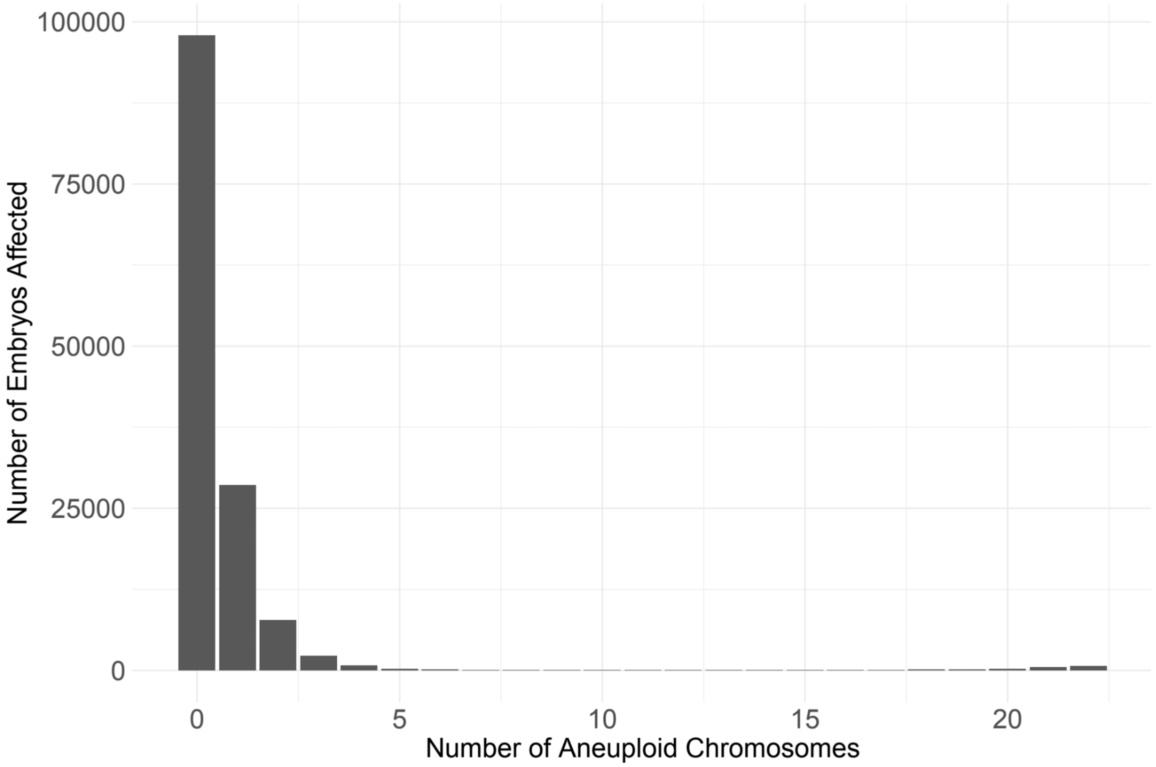


**Figure S4**. Distribution of aneuploid chromosomes across embryos. 70.25% of embryos in the dataset were found to be fully euploid, while 20.49% had an aneuploidy of a single chromosome, and 9.26% were affected by complex aneuploidy, including 1.309% overall that were subject to whole genome gain (1.078%) or loss/genome-wide uniparental isodisomy (0.231%). 577 embryos had between 6 and 18 aneuploid chromosomes.

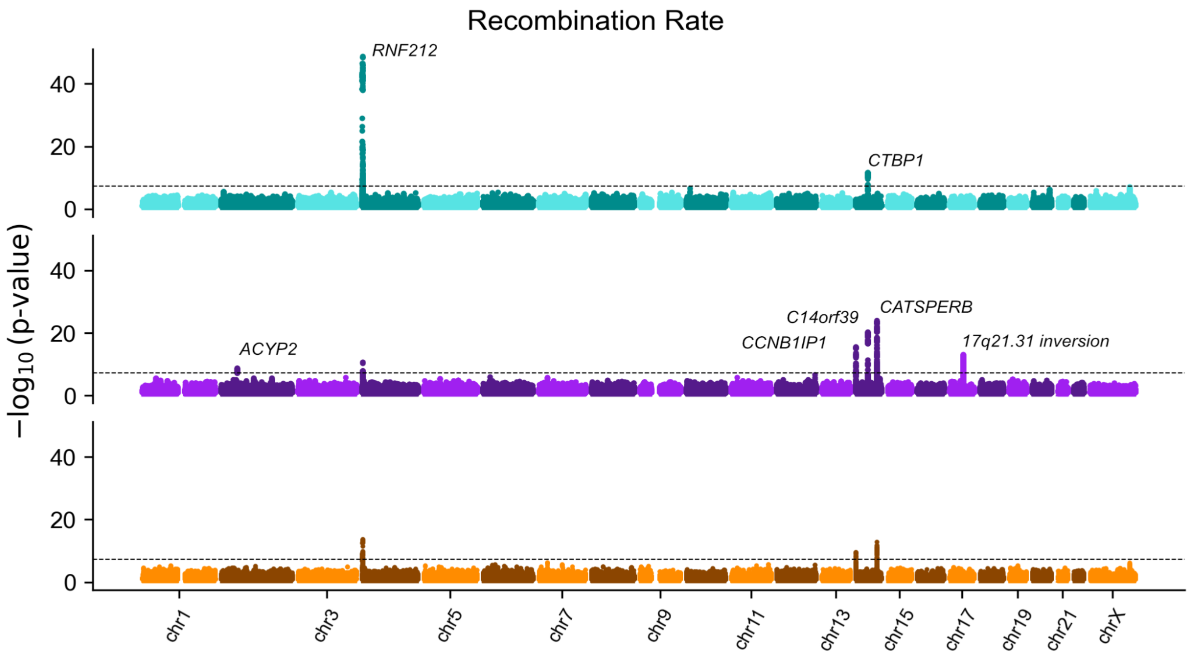


**Figure S5:** Manhattan plots for GWAS of recombination rate. Results per trait are stratified by paternal (cyan), maternal (purple) and joint (orange) analyses. Dashed line reflects the genome-wide significance threshold ($p < 5 \times 10^{-8}$). Physically nearest genes to lead GWAS variant per-peak are annotated per trait.

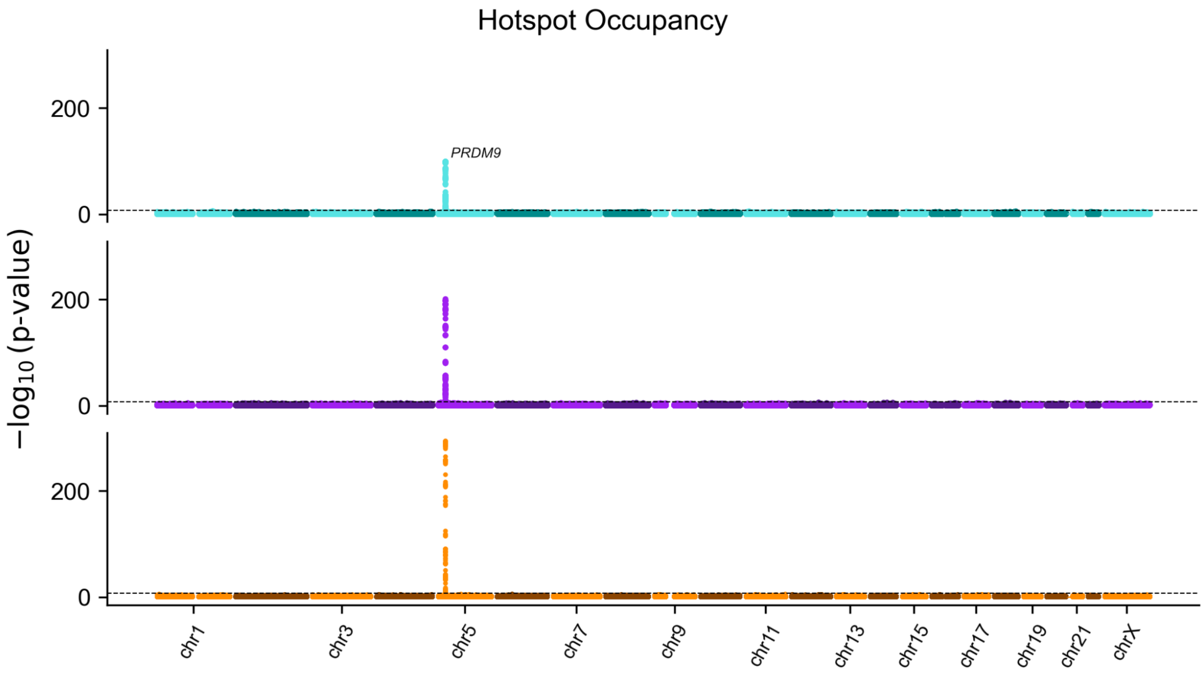


**Figure S6:** Manhattan plots for GWAS of recombination hotspot occupancy. Results per trait are stratified by paternal (cyan), maternal (purple) and joint (orange) analyses. Dashed line reflects the genome-wide significance threshold ($p < 5 \times 10^{-8}$). Physically nearest genes to lead GWAS variant per-peak are annotated per trait.

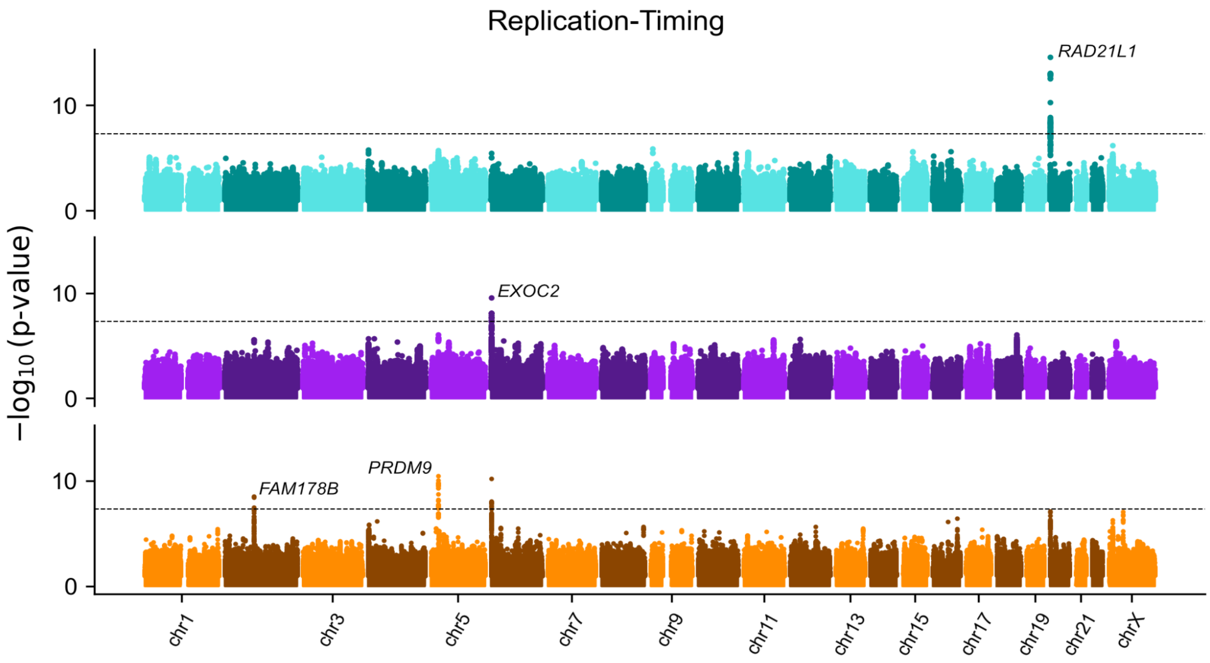


**Figure S7:** Manhattan plots for GWAS of estimated replication timing at hotspots. Results per trait are stratified by paternal (cyan), maternal (purple) and joint (orange) analyses. Dashed line reflects the genome-wide significance threshold ($p < 5 \times 10^{-8}$). Physically nearest genes to lead GWAS variant per-peak are annotated per trait.

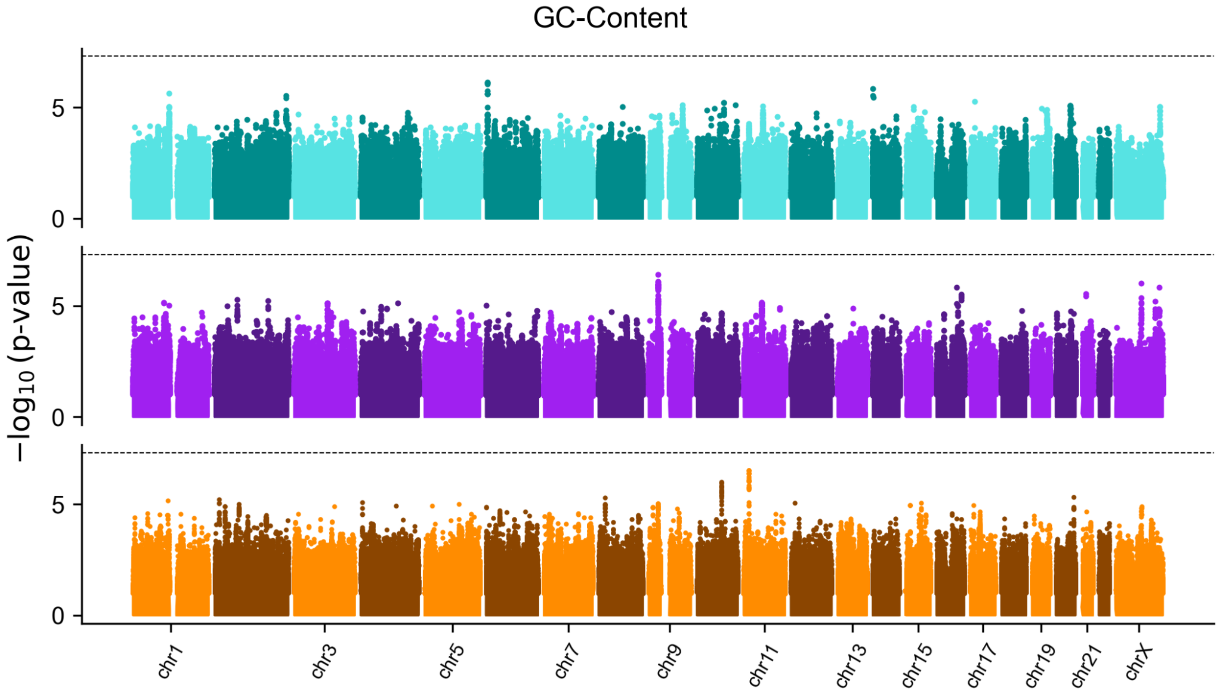


**Figure S8:** Manhattan plots for GWAS of GC content at crossover locations. Results per trait are stratified by paternal (cyan), maternal (purple) and joint (orange) analyses. Dashed line reflects the genome-wide significance threshold ($p < 5 \times 10^{-8}$). Physically nearest genes to lead GWAS variant per-peak are annotated per trait.

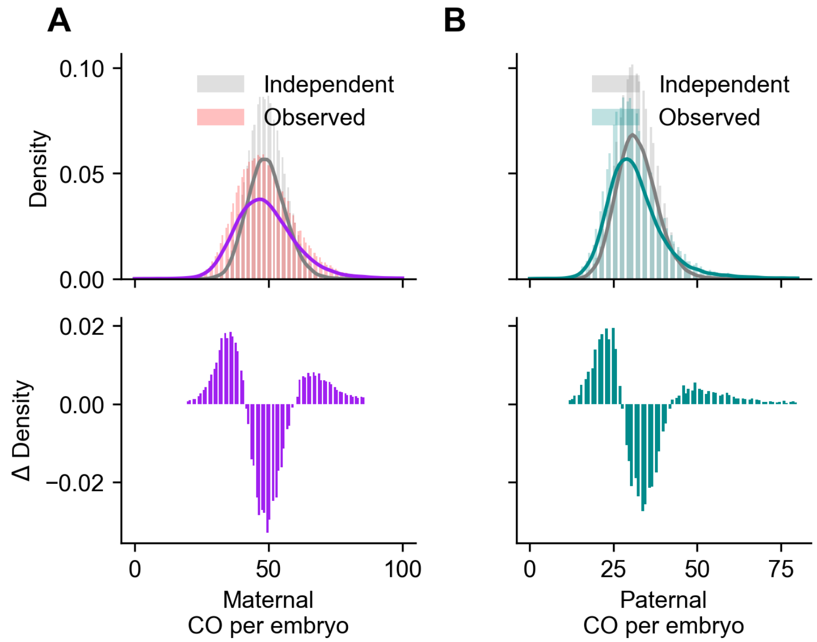


**Figure S9:** Comparison of observed crossover count (colored bars) against chromosomally independent simulations (gray bars) reflecting substantial overdispersion in crossovers due to positive covariance in (A) maternal and (B) paternal crossovers between chromosomes. Solid lines are kernel density estimates fit to the corresponding simulated and real crossover distributions.

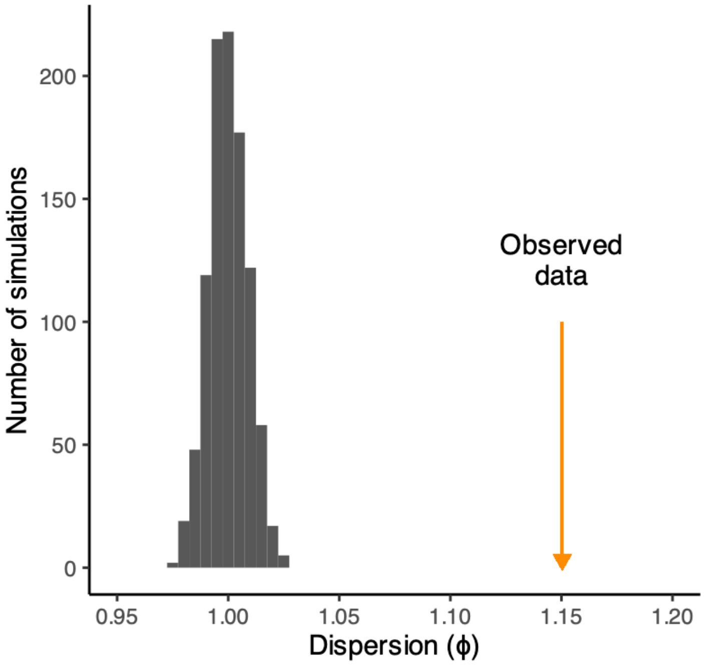


**Figure S10:** Dispersion parameters measured from data simulated under a binomial model (with no overdispersion), controlling for maternal age, compared to dispersion measured from the actual data. Dispersion is calculated as the sum of squared Pearson residuals, divided by the residual degrees of freedom.

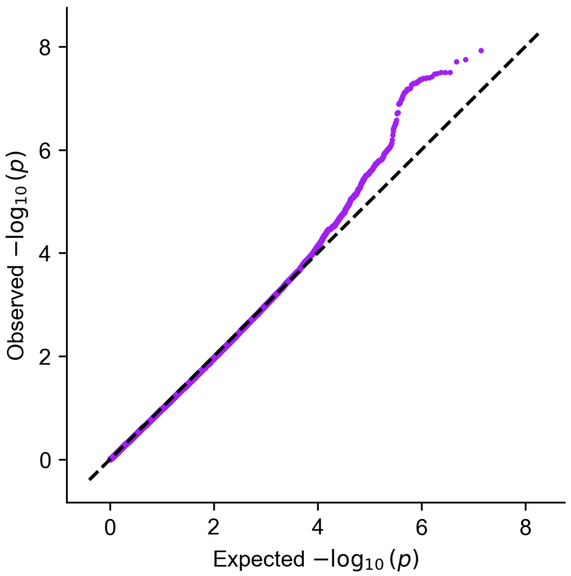


**Figure S11**. Quantile-quantile plot of GWAS for maternal meiotic aneuploidy.

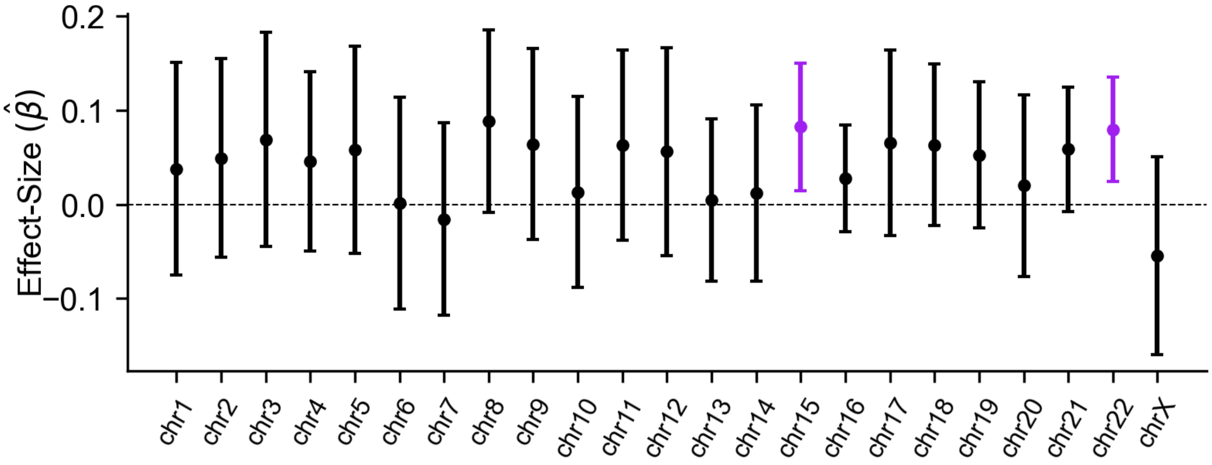


**Figure S12**. Marginal effect of rs6006737 on single-chromosome maternal meiotic aneuploidy. For each test, embryos aneuploid for the chromosome of interest are compared to euploid embryos, excluding those with aneuploidies affecting other chromosomes. While the effect-size estimates are positive for nearly all chromosomes, only chromosomes 15 and 22 are individually significant.

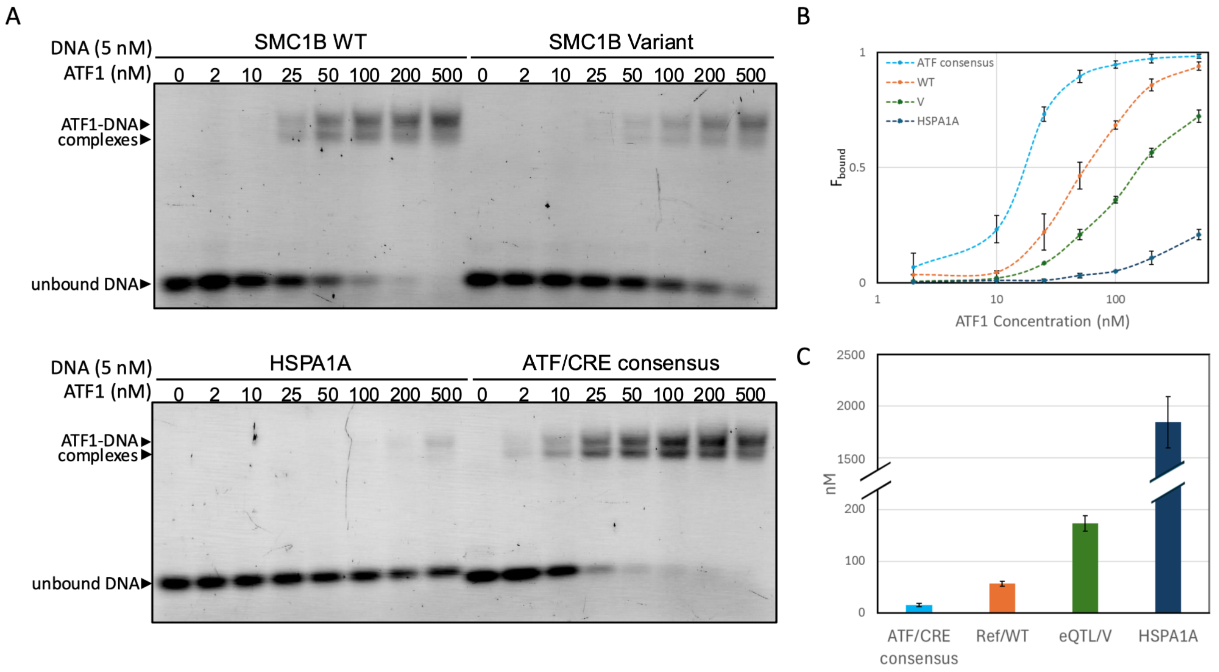


**Fig. S13**. (A) Representative gel scans of electrophoretic mobility shift assays for *SMC1B* WT and variant, ATF/CRE consensus sequence from the Epstein-Barr virus LMP1 gene promoter [124] as a positive control, and HSPA1A promoter sequence as a negative control. ATF1 is a basic-leucine zipper transcription factor; the two shifted ATF1-DNA complex bands indicate binding of ATF1 as a monomer or dimer. (B) The fraction of DNA bound as ATF1 concentration was titrated. Error bars indicate standard deviation from 2-4 replicates for each concentration. (C) Values for the $K_D$ obtained from curve-fit (see Methods). Error bars indicate standard deviation across four replicates.

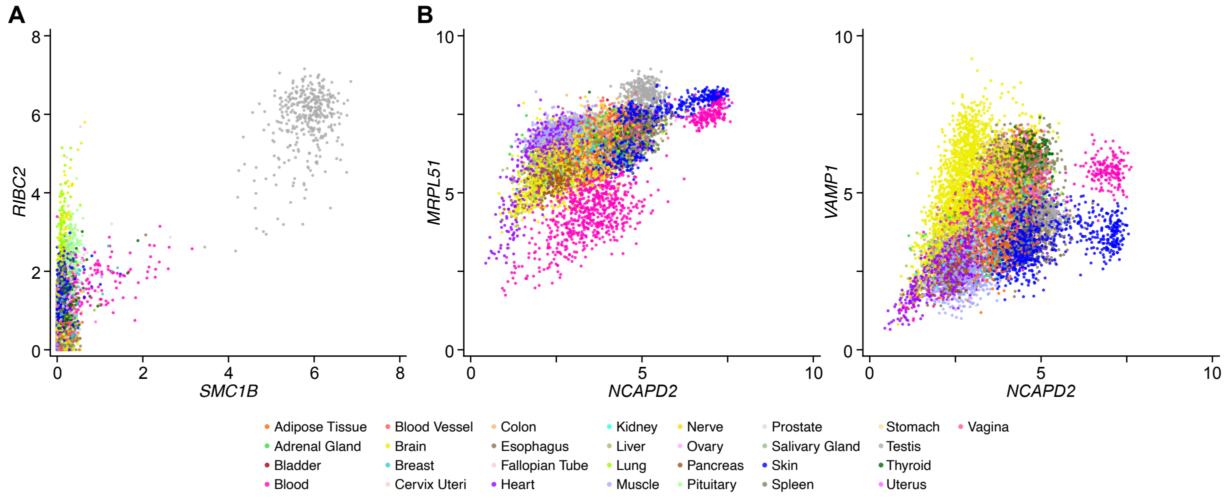


**Fig. S14**. Genes associated with meiotic machinery are co-expressed. Scatter plots compare $\log_2$(TPM+1) gene expression levels from GTEx v8 RNA-seq [41] at two aneuploidy TWAS loci with multiple associated genes. Expression of (A) *RIBC2* with *SMC1B* at the first TWAS locus and (B) *VAMP1* and *MRPL1* with *NCAPD2* at the second TWAS locus are highly correlated.

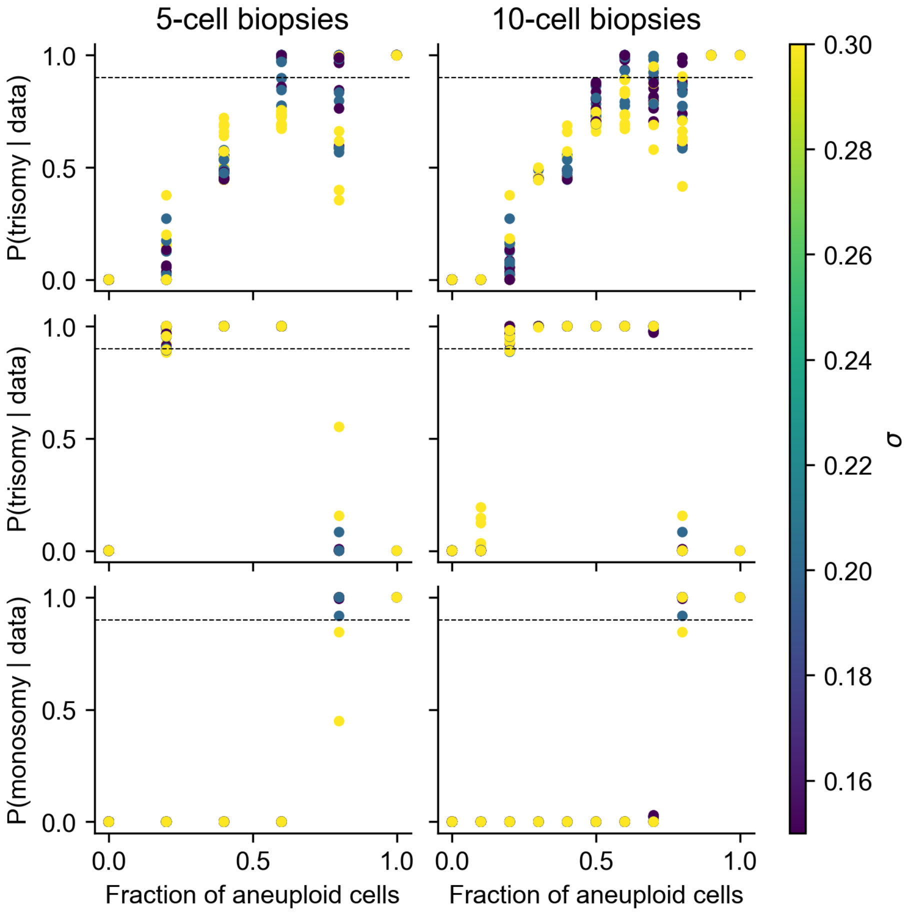


**Fig. S15.** Posterior probability of trisomy across 5-cell (first column) and 10-cell biopsies at different levels of genotyping array noise. The first row shows an increasing cell fraction of mosaic trisomies. The second and third rows show the cell-fractions of simulated monosomies and their effects on the estimated posterior probability of trisomy and monosomy respectively.

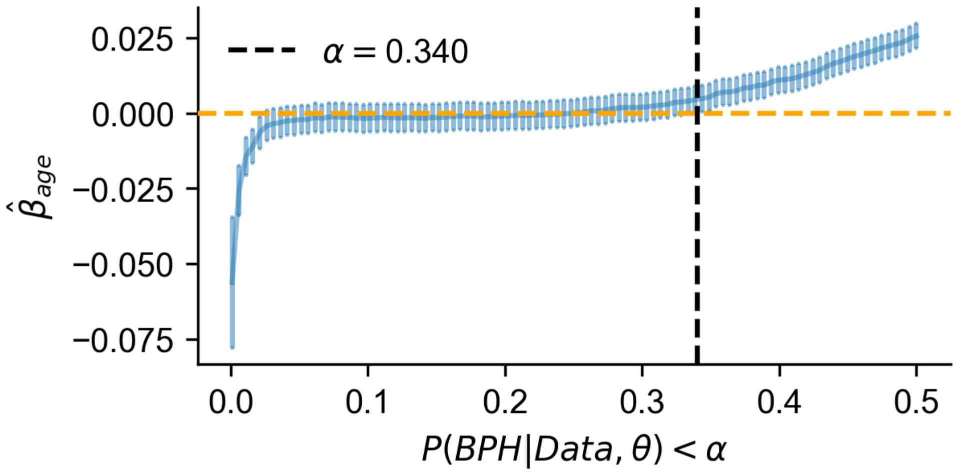


**Fig. S16.** Maternal age effect size from binomial regression model treating mosaic trisomies as those with a posterior probability of BPH < $\alpha$ in the full dataset. Error bars reflect two standard errors.

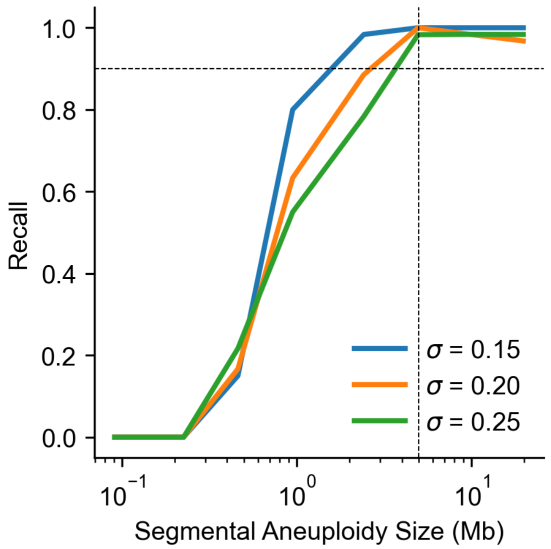


**Fig. S17.** Mean recall across 50 simulated segmental aneuploidies at different lengths of segmental gains/ losses (equally distributed) and across increasing noise thresholds (σ). Vertical line indicates five Mbp threshold used for identifying segmental aneuploidies.
